# Functional genomics provide key insights to improve the diagnostic yield of hereditary ataxia

**DOI:** 10.1101/2022.06.24.22276803

**Authors:** Zhongbo Chen, Arianna Tucci, Valentina Cipriani, Emil K. Gustavsson, Kristina Ibañez, Regina H. Reynolds, David Zhang, Letizia Vestito, Alejandro Cisterna García, Siddharth Sethi, Jonathan W. Brenton, Sonia García-Ruiz, Aine Fairbrother-Browne, Ana-Luisa Gil-Martinez, Genomics England Research Consortium, Nick Wood, John A. Hardy, Damian Smedley, Henry Houlden, Juan Botía, Mina Ryten

**Affiliations:** Department of Neurodegenerative Disease, Queen Square Institute of Neurology, University College London (UCL), London, WC1N 3BG, UK.; Department of Genetics and Genomic Medicine, Great Ormond Street Institute of Child Health, UCL, London, WC1N 1EH, UK.; NIHR Great Ormond Street Hospital Biomedical Research Centre, UCL, London, WC1N 1EH, UK.; William Harvey Research Institute, Queen Mary University of London, London, EC1M 6BQ, UK.; Genomics England, Queen Mary University of London, London, EC1M 6BQ, UK.; Ear Institute, UCL, London, WC1X 8EE, UK.; Department of Information and Communications Engineering, University of Murcia, 30003 Murcia, Spain.; Astex Pharmaceuticals, 436 Cambridge Science Park, Cambridge, CB4 0QA, UK.; Department of Medical and Molecular Genetics, School of Basic and Medical Biosciences, King’s College London, London, WC2R 2LS, UK.; Department of Clinical and Movement Neurosciences, Queen Square Institute of Neurology, UCL, London, WC1N 3BG, UK.; Reta Lila Weston Institute, Queen Square Institute of Neurology, UCL, London, WC1N 3BG, UK.; UK Dementia Research Institute at UCL, London, WC1E 6BT, UK.; NIHR University College London Hospitals Biomedical Research Centre, London, W1T 7DN, UK.; Institute for Advanced Study, The Hong Kong University of Science and Technology, Hong Kong SAR, China.; Department of Neuromuscular Disease, Queen Square Institute of Neurology, UCL, London, WC1N 3BG, UK.

**Keywords:** hereditary ataxia, functional genomics, transcriptomics, repeat expansion disorders, rare disease

## Abstract

Improvements in functional genomic annotation have led to a critical mass of neurogenetic discoveries. This is exemplified in hereditary ataxia, a heterogeneous group of disorders characterised by incoordination from cerebellar dysfunction. Associated pathogenic variants in more than 300 genes have been described, leading to a detailed genetic classification partitioned by age-of-onset. Despite these advances, up to 75% of patients with ataxia remain molecularly undiagnosed even following whole genome sequencing, as exemplified in the 100,000 Genomes Project.

This study aimed to understand whether we can improve our knowledge of the genetic architecture of hereditary ataxia by leveraging functional genomic annotations, and as a result, generate insights that raise the diagnostic yield.

To achieve these aims, we used publicly-available multi-omics data to generate 294 genic features, capturing information relating to a gene’s structure, genetic variation, tissue-specific, cell-type-specific and temporal expression, as well as protein products of a gene. We studied these features across genes typically causing childhood-onset, adult-onset or both types of disease first individually, then collectively. This led to the generation of testable hypotheses which we investigated using whole genome sequencing data from 2,322 individuals presenting with ataxia and 6,387 non-neurological probands in the 100,000 Genomes Project.

Using this approach, we demonstrated a surprisingly high short tandem repeat (STR) density within childhood-onset genes suggesting that we may be missing pathogenic repeat expansions within this cohort. This was verified in both childhood- and adult-onset ataxia patients from the 100,000 Genomes Project who were unexpectedly found to have a trend for higher repeat sizes even at naturally-occurring STRs in known ataxia genes, implying a role for STRs in pathogenesis. Using unsupervised analysis, we found significant similarities in genomic annotation across the gene panels, which suggested adult- and childhood-onset patients should be screened using a common diagnostic gene set. We tested this within the 100,000 Genomes Project by assessing the burden of pathogenic variants among childhood-onset genes in adult-onset patients and vice versa. This demonstrated a significantly higher burden of rare, potentially pathogenic variants in conventional childhood-onset genes among individuals with adult-onset ataxia.

Our analysis has implications for the current clinical practice in genetic testing for hereditary ataxia. We suggest that the diagnostic rate for hereditary ataxia could be increased by removing the age-of-onset partition, and through a modified screening for repeat expansions in naturally-occurring STRs within known genes, in effect treating these regions as candidate pathogenic loci.

## Introduction

Over the last two decades, there has been significant progress in the diagnosis of neurogenetic diseases.^1–3^ Despite this, approximately half of patients presenting with a probable genetic cause for a neurological disorder remain undiagnosed,^1, 4^ with the most clinically and genetically heterogeneous disorders presenting the greatest challenge.^5, 6^ One such archetypal heterogeneous neurogenetic condition is hereditary ataxia. These are a group of neurodegenerative disorders characterised by the clinical syndrome of progressive incoordination due to cerebellar dysfunction^7, 8^ with a prevalence of approximately 1.5 to 4.9 per 100,000 persons.^9^ To date, variants in more than 300 genes have been discovered to be associated with ataxia taking us away from Greenfield’s patho-anatomical and Harding’s clinico-genetic classifications.^10^ Despite this shift towards a detailed molecular classification, diagnostic rates remain relatively low.^8, 11, 12^ Even when using whole genome sequencing (WGS) in a highly-selective cohort in the Genomics England 100,000 Genomes Project, the diagnostic yield for hereditary ataxia was only 21% among singletons and 32% in family trios.^13^ This could be explained both by the existence of as yet undiscovered causative genes and the incomplete screening of known genes in the appropriate patients.

The current genetic evaluation strategy in clinical practice for hereditary ataxia involves partitioning patients by age-of-onset; in the UK, this is employed by the 100,000 Genomes Project^14^ and NHS England National Genomic Test Directory (https://www.england.nhs.uk/publication/national-genomic-test-directories/). Diagnostic- grade panels of genes for ataxia are divided into childhood-onset, adult-onset and cerebellar hypoplasia categories.^14^ This is also reflected in the Childhood Ataxia and Cerebellar Group of the European Paediatric Neurology Society guidelines, which suggests a specific evaluation algorithm (including genetic testing) for early-onset ataxia.^15^ In practice, the existence of these separate panels means that patients with adult-onset ataxia are seldom screened for genes typically associated with childhood-onset disorders such as Joubert syndrome and childhood-onset patients are rarely screened for adult-onset variants such as pathogenic repeat expansions typically associated with some late-onset spinocerebellar ataxia (SCA).^16^ It is difficult to assess with confidence whether this age-based classification is justified. Most genetic variants associated with ataxia have only recently been recognised and consequently reported in just a handful of cases.^6^ This makes it challenging to determine whether this age-of-onset division reflects the true biology of disease or arises from presentation bias.

With increasing quantities of publicly-available functional multi-omic annotations that assign biological meaning to genomic regions, this has become a tractable question.^17^ Through the application of multi-omics technologies and computational tools, it is possible to produce increasingly precise and granular annotations that operate at both a tissue- and cell-type-specific level and provide insights into the shared biology of disease-associated genes.^18, 19^ This has been key for neurological diseases and importantly, advances in this field have already improved our interpretation of gene-phenotype associations and driven genetic discovery.^18, 20^ Therefore, we questioned whether the improvements in the breadth and depth of functional multi-omic annotations would enable us to identify commonalities and hidden biological relationships among genes causing ataxia. Furthermore, we investigated whether this information could be used to improve our understanding of the underlying genetic architecture of these diseases and potentially inform novel testing strategies that could increase diagnostic yield.

In this study, we leveraged the critical mass of genes discovered in ataxia together with a curated set of approximately 300 multi-omic genic features to determine whether ataxia genes can be characterised on a genomic level to identify hidden patterns that explain their biology. We also used this analysis to determine whether the clinical division of ataxia by age-of-onset is reflected in genomic annotations with implications for the current diagnostic strategy. Finally, we assessed the potential of alternative testing strategies using WGS of 2,322 probands presenting with ataxia as either a primary or secondary symptom recruited in the 100,000 Genomes Project. Using this approach, we gained further insight into the pathogenic mechanisms underlying hereditary ataxia and highlighted potential bottlenecks to diagnosis.

## Materials and methods

### Defining a list of genes associated with hereditary ataxia

In order to identify genes known to be associated with hereditary ataxia across all ages, we used three resources: (i) Genomics England PanelApp, a publicly-available crowdsourced tool to standardise gene panels^14^; (ii) Hereditary Ataxia GeneReviews, a regularly updated international point-of-care resource on inherited medical conditions^7^; and (iii) the Online Mendelian Inheritance in Man (OMIM) database.^21^ We extracted “*green*”, diagnostic-grade genes from PanelApp considering the four gene panels of cerebellar hypoplasia (v.1.41), adult-onset hereditary ataxia (v.2.8), childhood-onset ataxia (v.6.22) and hereditary ataxia (v.1.2.05). By combining the resources available in PanelApp, GeneReviews and the OMIM database, we identified a total of 318 unique genes (workflow shown in **Fig. 1**). We noted that there were discrepancies between the resources in the expected age-of-onset. For example, *SCYL1* was classified in PanelApp as adult-onset but has been described as a childhood-onset disorder (spinocerebellar ataxia autosomal recessive 21. MIM:616719).^21^ Thus, to standardise information for genes of interest, we extracted typical age-of-onset data in an automated manner from OMIM “Text” section (http://api.omim.org)^21^, and also manually curated information from OMIM “Clinical Synopsis” that reviews reported cases. Thus, the 318 genes identified were classified into: (i) adult-onset (*N*=23, typical age-of-onset ≥18 years); (ii) childhood-onset (*N*=213, typical age-of-onset <18 years); and (iii) overlap-onset genes (*N*=82, causing both childhood- and adult-onset disease) (**Supplementary Table 1**). Given that we sought to differentiate between genes associated with ataxia and genes not known to cause ataxia, we defined the set of remaining protein-coding genes as controls (*N*=17,323, Ensembl v.72).^22^

**Figure 1.**
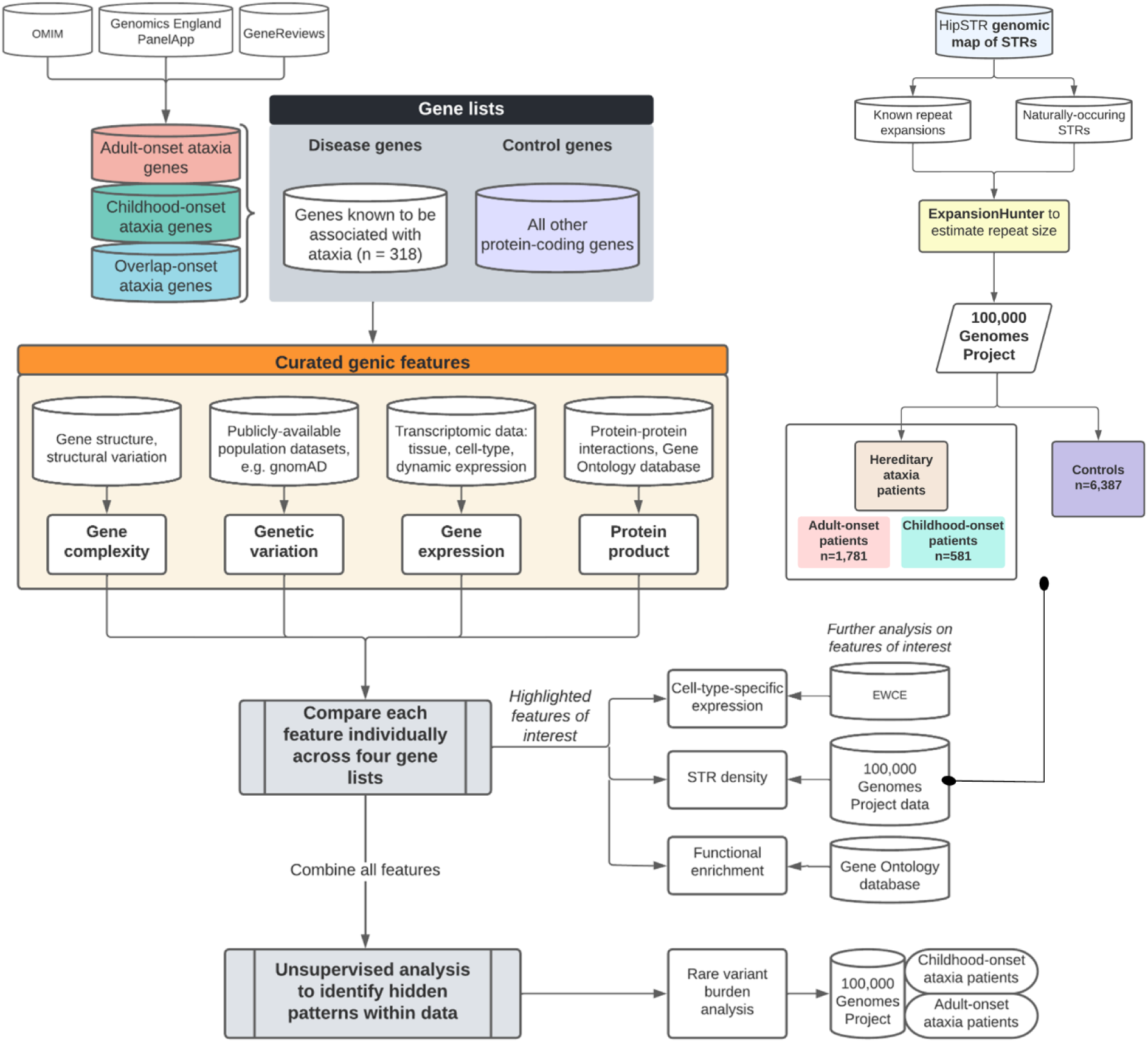
Overall workflow of study. Genic information is captured across categories of genetic variation, gene structure/complexity, gene expression and co-expression and protein- product of a gene, and compared across the four gene lists of: (i) adult-onset ataxia genes; (ii) childhood-onset ataxia genes; (iii) overlap-onset ataxia genes, defined as those associated with both childhood- and adult-onset, when mutated; (iv) other protein-coding genes not known to cause ataxia (control “not ataxia” genes). The gene lists were extracted primarily from Genomics England PanelApp, but also GeneReviews and OMIM. The age-of-onset definition was derived primarily from OMIM to reduce bias. Genic features were first compared individually across the four gene lists then combined together through unsupervised clustering analysis. Individual genic features were also highlighted and put through further analyses including expression-weighted cell-type enrichment (EWCE) for cell-type-specific expression and functional gene ontology (GO) enrichment. Further verification of the results from functional genomic annotation were verified in whole genome sequencing data of patients with ataxia recruited to the 100,000 Genomes Project through rare variant burden analysis and short tandem repeat (STR) analysis.

### Extracting clinical phenotype information

To capture genic information about the clinically-heterogeneous phenotypes associated with hereditary ataxia genes, we used data provided within the OMIM catalogue (http://api.omim.org)^21^ and the Human Phenotype Ontology (HPO) database (http://compbio.charite.de/jenkins/job/nhpo.annotations/lastStableBuild/artifact/util/annotation/genes_to_phenotype.txt, Build 1271)^23^ as the latter also incorporated additional information from Orphanet.^24^ We extracted the HPO and OMIM terms associated with each gene.

### Genic feature generation

We curated a total of 294 genic features leveraging publicly-available multi-omics datasets capable of providing genome-wide information. The genic features captured information in four main categories relating to: (i) gene structure and complexity; (ii) genetic variation including evolutionary features; (iii) gene expression and co-expression; (iv) protein product of a gene. A full list of genic features with their corresponding source is provided in **Supplementary Table 2**.

#### Gene structure and complexity

We extracted information relating to gene structure from Ensembl v.72.^25^ This included: gene length; number of unique transcripts within a gene; number of exon-exon junctions and the gene’s GC content among others. Specific to the pathogenic mechanisms of ataxia, we used resources generated through the application of Tandem Repeats Finder^26^ to the human reference genome (GRCh38)^27^ in HipSTR (https://github.com/HipSTR-Tool/HipSTR-references) to create a gene-based metric of short tandem repeat (STR) density, size, number of nucleotides within each STR, and location of STRs as annotated by Ensembl v.72.^22^ This was complemented by extraction of information on STRs associated with expression of nearby genes (eSTRs).^28^ We generated additional annotations to reflect the presence of other repetitive elements using the RepeatMasker reference panel (http://www.repeatmasker.org).^29,30^ These included short interspersed nuclear (SINE)/Alu elements, Retroposon/SVAs, and long interspersed nuclear (LINE)/L1 elements (full list in **Supplementary Table 2**).

#### Genetic variation including evolutionary information

Measures of genetic variation were collated from existing large population databases and the related resources.^31^ We used LoFTool (gene intolerance score based on loss-of-function (LoF) variants)^32^, EvoTol (measures a gene’s intolerance to mutation using evolutionary conservation of protein sequences)^33^, RVIS (intolerance scoring system of a gene’s functional variation)^34^, gnomadpLI (LoF score from gnomAD such that a pLI closer to 1 indicates that the gene or transcript cannot tolerate protein-truncating variation)^31^ among other metrics. We also derived features to capture evolutionary information about a gene. Using the phastCons20 score, a measure of inter-species conservation between primates^35^ together with context dependent tolerance score (CDTS), a measure of intra-species constraint^36^, we generated the genic density of constrained, non-conserved genomic regions (CNCRs), which represent human-lineage-specific element annotation.^37^

#### Gene expression and co-expression

We leveraged publicly-available transcriptomic data to capture information on tissue-specific, cell-type-specific and temporally-relevant expression. Using temporal expression data generated from RNA-sequencing of human organ development over 23 timepoints from four weeks post-conception to the sixth decade of life, we obtained a measure of developmental gene expression (developmentally-dynamic expression pleiotropy index (0–1) with a value of 1 indicating the most dynamically- repressed expression).^38^ Using data from 47 Genotype-Tissue Expression Project (GTEx) tissues, gene expression data were filtered for genes with >0.1 Reads Per Kilobase Million (RPKM) and corrected for batch effects, age, sex and RNA integrity number using ComBat.^39^ Residuals of these linear regression models were used to calculate tissue-specific expression and to construct gene co-expression networks (WGCNA) for each tissue (R package G2PML).^25^ A gene was defined as having tissue-specific expression if expression in that tissue was five-fold higher than the mean across all tissues. For each gene in a network, the Module Membership (MM) for a gene was defined as the correlation of its residual gene expression and the eigengene of the module to which it belonged^25^. A gene had tissue-specific MM at 3.5-fold higher than across all other GTEx tissues. For cell-type-specific expression, we used single cell RNA-sequencing data from mice.^40^ We generated features incorporating information from specificity matrices of expression data.^41^ We used all level 2 and level 3 cell types within and outside the CNS. We then focused on cerebellar-specific expression (CNS superset level 2).^40^

#### Curated data on the protein product of a gene and its function

The Gene Ontology (GO)^42, 43^ and STRING databases were used to obtain information on the function of a gene’s protein product and the number of its protein-protein interactions.^44^ We used g:Profiler^45^ for GO enrichment analysis, with the g:SCS multiple testing correction method. In addition, we used the Full Spectrum of Intolerance to Loss-of-function (FUSIL) categorisation of genes, which was based on both viability and phenotyping screens performed on knock-out mice, and essentiality screens carried out on human cell lines.^46^ This provided five gene categories relating to the essentiality of their function: cellularly lethal (CL); developmentally lethal (DL); subviable (SV); viable with significant phenotype/s (VP) or viable with no significant phenotypes detected (VN).^46^

### Statistical analysis

For each feature generated, we compared the feature across the four gene lists, namely adult-onset, childhood-onset, overlap-onset ataxia and control genes. For continuous variables, we used Wilcoxon rank sum test to compare the means of the metrics between the groups, taking a 2-tailed p-value < 0.05 as significant. For categorical variables, we used chi-squared tests to assess statistically significant (*P<*0.05) differences between the distributions of two groups. We have outlined statistical analyses of other methods within the relevant sections. All analyses were carried out in R (v.6.1).

### Expression-weighted cell-type enrichment

Expression-weighted cell-type enrichment (EWCE) was used to determine whether ataxia genes have higher expression within particular brain-related cell types than would be expected by chance (https://github.com/NathanSkene/EWCE).41 We used the adult-, childhood- and overlap-onset ataxia gene lists as input with specificity matrices calculated for level 2/3 cell types,^40^ as well as level 2 cell types from the superset containing cerebellar-specific cell types. We controlled for transcript length and GC-content in the bootstrap lists where EWCE was run with 10,000 bootstrap replicates. Genes without a 1:1 mouse:human ortholog were excluded. *P*-values for multiple testing were corrected using the Benjamini-Hochberg method over all cell types and the three gene sets. At the cerebellar-specific cell type level, *P*-values were only corrected for the number of gene sets given the granularity of cell-types at this level. To assess the contribution of specific genes to cell-type-specific expression, we obtained the mean expression for the gene of interest within the cell type divided by its expression across all cell types.^41^

### Unsupervised analysis of all features

In order to compare the utility of all features in classifying ataxia genes, first, we used recursive feature elimination (caret R package^47^) to remove redundant features, defined as those with Pearson’s correlation >0.9 between two features (visualised using corrplot package^48^). This approach also helped to account for the imbalance in gene list sizes by taking a random sample (50%) of disease genes and an equal number of bootstrap-selected control genes from the larger control set. The recursive feature elimination then took a random subset of input features based on feature importance and assessed the number of features with the highest accuracy as defined by Cohen’s ĸ statistic. It then fitted a generalised linear model for the best set of features. The feature elimination operates in a *k*- fold cross-validation manner to improve the accuracy of the fit of the model for unseen samples. We extracted the minimum proportion of times that a feature was selected as most relevant out of the total number of iterations in the repeat variable (parameter *r*). In this case, we used features that had an *r* of ≥0.8, indicating that they appeared more than 40 times out of the 50 repetitions performed. Taking these selected features, we used uniform manifold approximation and projection (UMAP) to investigate any hidden patterns within the data defined using these salient features.^49^

### Validation within 100,000 Genomes Project participants

#### 1. Genetic variant burden testing

To test whether adult- and childhood-onset hereditary ataxia patients should be screened for mutations using a common gene set, a case-control gene burden analysis was adapted from an analytical framework used within the rare disease component of the 100,000 Genomes Project.^13^ First, cases were defined as all probands recruited under the clinical indications: “hereditary ataxia”, “cerebellar hypoplasia” or “pontine tegmental cap dysplasia”. For the corresponding control group, we used all other probands aged ≥40 years at recruitment excluding individuals recruited under relevant neurological or related disease categories (*N=*3720) (exclusion criteria for controls in **Supplementary Table 3**). We used the recorded age-of-onset, where available, or age at recruitment as proxy. Cases were defined as childhood-onset (<18 years) (*N=*306) and adult-onset (≥18 years) (*N=*816). Using these definitions, four different combinations of rare genetic variant burden analyses were performed as summarised in **Supplementary Table 4**: (1) childhood-onset genes in adult-onset cases *vs.* in controls; (2) adult-onset genes in childhood-onset cases *vs*. in controls; (3) overlap-onset genes in adult-onset cases *vs.* in controls; (4) overlap-onset genes in childhood-onset cases *vs.* in controls, with the latter two analyses used for controlled comparison as we would expect the find overlap-onset variants in both childhood- and adult-onset cases.

The sets of rare variants for the gene burden analyses were obtained running Exomiser^50^ on all probands’ WGS data to filter coding variants that are rare (minor allele frequency_<_0.1%, for dominant, and <1%, for recessive variants in gnomAD^31^ v2.1.1 and v3.1.1 as well as within the local 100,000 Genomes Project cohort and segregated with disease status (where family information was available). Gene-based enrichment of rare variants in cases was assessed using one-sided Fisher’s exact test under four scenarios: (1) enrichment of rare, predicted LoF variants, (2) enrichment of rare, predicted pathogenic variants (Exomiser variant score_>0.8), (3) enrichment of rare, predicted pathogenic variants in a constrained coding region, (4) enrichment of rare, *de novo* variants. For the latter, only trios or larger families where *de novo* calling was possible were considered. To maintain statistical validity and power, the analysis was limited to those disease-gene associations where relevant variants in the gene were seen in at least four probands over the entire cohort of cases and controls. Benjamini-Hochberg method was used to correct for multiple testing; an overall FDR *P* threshold of 0.10 was used for claiming significant gene-disease associations accounting for the total number of case-control gene burden tests under all four scenarios, i.e., 360.

To support this, we then reviewed these patients with ataxia for apparent incongruities between the age of disease onset and age-of-onset category associated with the diagnostic pathogenic variant on the final formal genetic report issued.

### STR sizes in participants with ataxia

We investigated whether naturally-occurring STRs within the 318 known ataxia genes harboured differences in repeat size distributions between individuals presenting with ataxia and controls recruited in the 100,000 Genomes Project. Ataxia cases were any probands presenting with ataxia, cerebellar hypoplasia or pontine tegmental cap dysplasia either within the enrolled disease group or as an HPO term (i.e., ataxia as a primary symptom or as part of a more complex syndrome) (adult-onset *N*=1760, childhood-onset *N*=553) and non-neurological controls aged ≥40 years (*N*=6078) from all recruited probands defined as per **Supplementary Table 4**. STR genotyping was performed using ExpansionHunter v.3.1.2^51^ using methods as previously described^16^ at HipSTR reference loci for naturally-occurring trinucleotides, tetranucleotides, pentanucleotides and hexanucleotides located in the exons or 5’UTRs of the 318 genes across all cases and controls.^27^ Using this definition, we studied a total of 197 STRs across 107 unique genes with seven being known pathogenic repeats. We then ranked the maximum repeat size for each STR and partitioned the repeat sizes into bins for controls and for cases. Taking the top 1% repeat sizes in controls and the top 1% in ataxia cases, partitioned by age-of-onset, we compared the mean differences in the repeat sizes. We applied the same testing strategy to STRs known to cause repeat expansion disorders, and which would be expected to have a higher repeat size in ataxia cases compared to controls.

### Data availability

The authors confirm that the data supporting the findings of this study are available within the article and its supplementary material. Sources for publicly-available data used for generating the genic features are shown in **Supplementary Table 2**.

## Results

### Childhood- and adult-onset ataxia differ in pathogenic variant type and phenotypic presentations

We analysed 318 genes known to cause hereditary ataxia when mutated, classified as: (i) childhood-onset (*N*=213), (ii) adult-onset (*N*=23), and (iii) overlap-onset genes (associated with either childhood- or adult-onset disease, *N*=82). We found that childhood-onset genes showed a higher proportion of biallelic autosomal inheritance compared with adult-onset (chi-squared *P*=2.329×10^-7^) and overlap-onset genes (*P*=6.094×10^-7^). In contrast, a significantly higher proportion of pathogenic repeat expansions cause adult-onset (34.8%) compared with childhood-onset disease (0.5%) (chi-squared *P*=3.205×10^-14^) (**Supplementary Fig. 1**). Using known phenotypic associations captured within OMIM and HPO^23^ showed that childhood-onset ataxia genes had the highest mean number of associated HPO terms per gene (**Fig. 2A**) and affected a significantly larger number of distinct body systems compared with adult-onset (Wilcoxon rank sum *P*=5.400×10^-4^) and overlap-onset genes (*P*=1.800×10^-4^) (**Fig. 2B, Supplementary Fig. 1C**). In summary, we found that childhood-onset ataxia genes were more likely to be autosomal recessive, less likely to be associated with repeat expansion disorders and tended to manifest in multiple systems compared with adult-onset genes.

**Figure 2.**
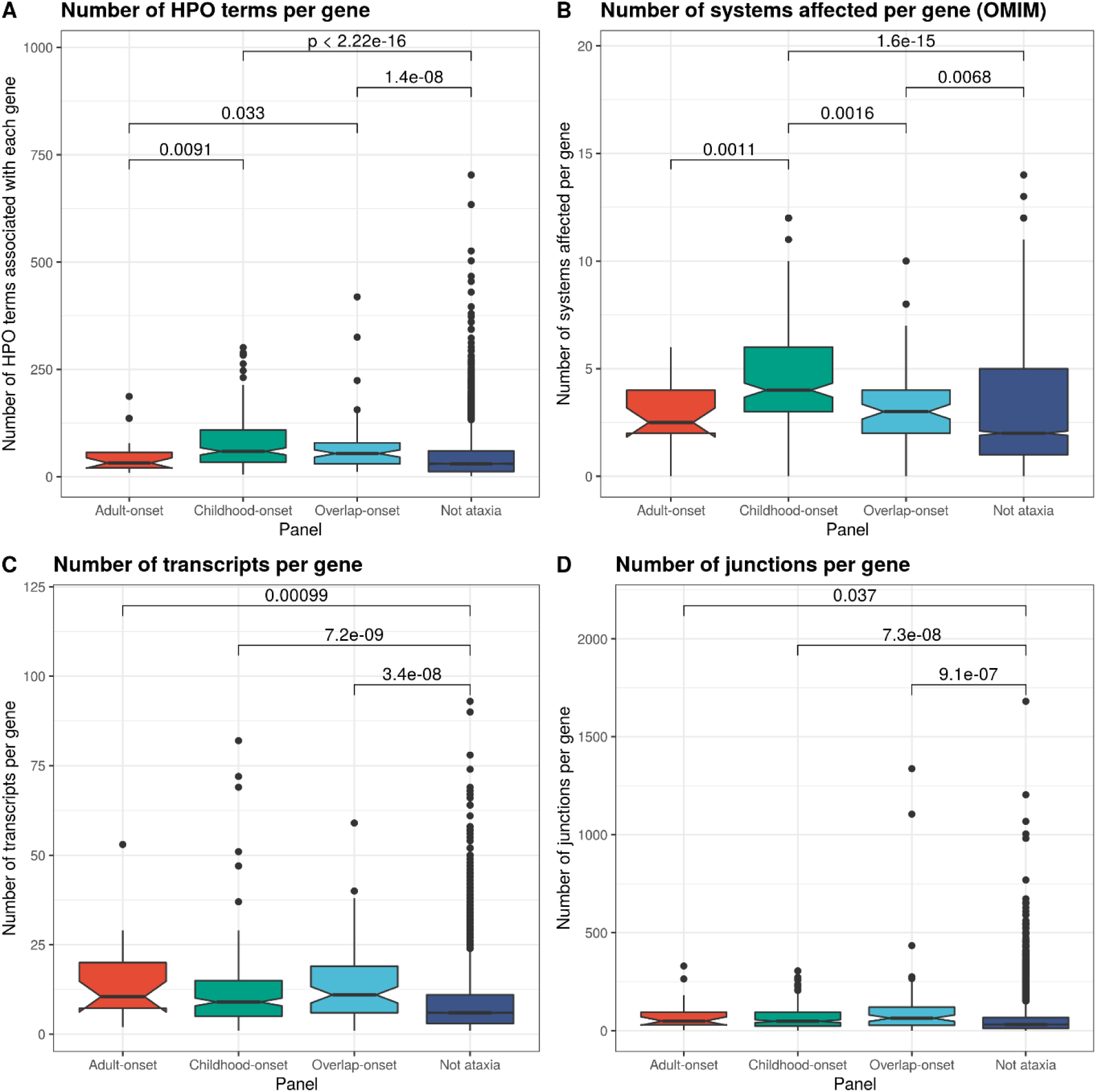
Comparison of phenotypes associated with genes as annotated by Human Phenotype Ontology (HPO) and Online Mendelian Inheritance in Man (OMIM) and gene complexity features between different gene panels. The number of known HPO terms associated with each gene is shown in **A.** The number of body systems affected associated with each gene as annotated by OMIM is shown in **B.** The number of transcripts of each known gene as annotated by Ensembl v.72 is shown in **C.** The number of annotated junctions within each gene as annotated by Ensembl v.72 is shown in **D**. Only significant Wilcoxon rank sum p-values (<0.05) are given for pairwise comparisons above the square brackets.

### Ataxia genes contain an increased density of repetitive elements

We leveraged the increasing availability of functional genomic annotation to expand our understanding of genes associated with hereditary ataxia. This analysis was performed by comparing each individual genic feature from our collation of ∼300 functional multi-omic annotations across the four gene lists (*P*-values of all comparisons in **Supplementary Table 5)**. We began by focusing on measures of gene complexity and structure. Using this approach, we saw increased overall complexity amongst genes associated with ataxia compared to the control set. Ataxia genes harbour more transcripts and junctions per gene, suggesting that splicing variants could contribute to pathogenesis (**Fig. 2C and D**). However, we noted that gene complexity was a feature of all disease gene sets and did not distinguish between childhood-, adult- and overlap-onset ataxia genes (**Supplementary Fig. 2**).

Next, we expanded our analysis to consider the impact of STRs on the complexity of ataxia gene structure. Although pathogenic STR expansions are already known to be an important disease-causing mechanism for adult-onset hereditary ataxia^52^ (**Supplementary Fig. 1, Fig. 3A**), we questioned whether a higher genic density of non-pathogenic STRs could also be a distinguishing property of ataxia genes. Generating a genomic map of STR elements,^27^ we found that the majority of intragenic STRs and eSTRs reside in the intron (95.13% and 94.43% respectively) (**Fig. 3A**). Furthermore, we noted the existence of 1,143 naturally-occurring intragenic CAG repeats which could be considered candidate loci for further interrogation in an unsolved cohort (**Fig. 3A**). Focusing on ataxia genes, we found that this gene set harboured a higher number of STRs per gene (median 34.5 and 27 STRs per adult- and childhood-onset gene respectively) than control genes (median 16 STRs per gene) (**Fig. 3B**). Surprisingly, this was evident when comparing childhood-onset genes to non-ataxia genes (*P=*0.008), although only adult-onset genes had a significantly higher trinucleotide repeat density than controls (**Fig. 3C**). This trend extended to eSTRs, defined as STRs associated with variable expression of nearby genes proportional to their repeat length.^28^ We found that childhood-onset genes had a higher number of associated eSTRs compared to control genes (*P*=7.300×10^-4^) (**Fig. 3D**), and that these eSTRs were detected in a higher number of tissues compared with control genes (*P*: 4.900×10^-5^) (**Fig. 3E**).

**Figure 3.**
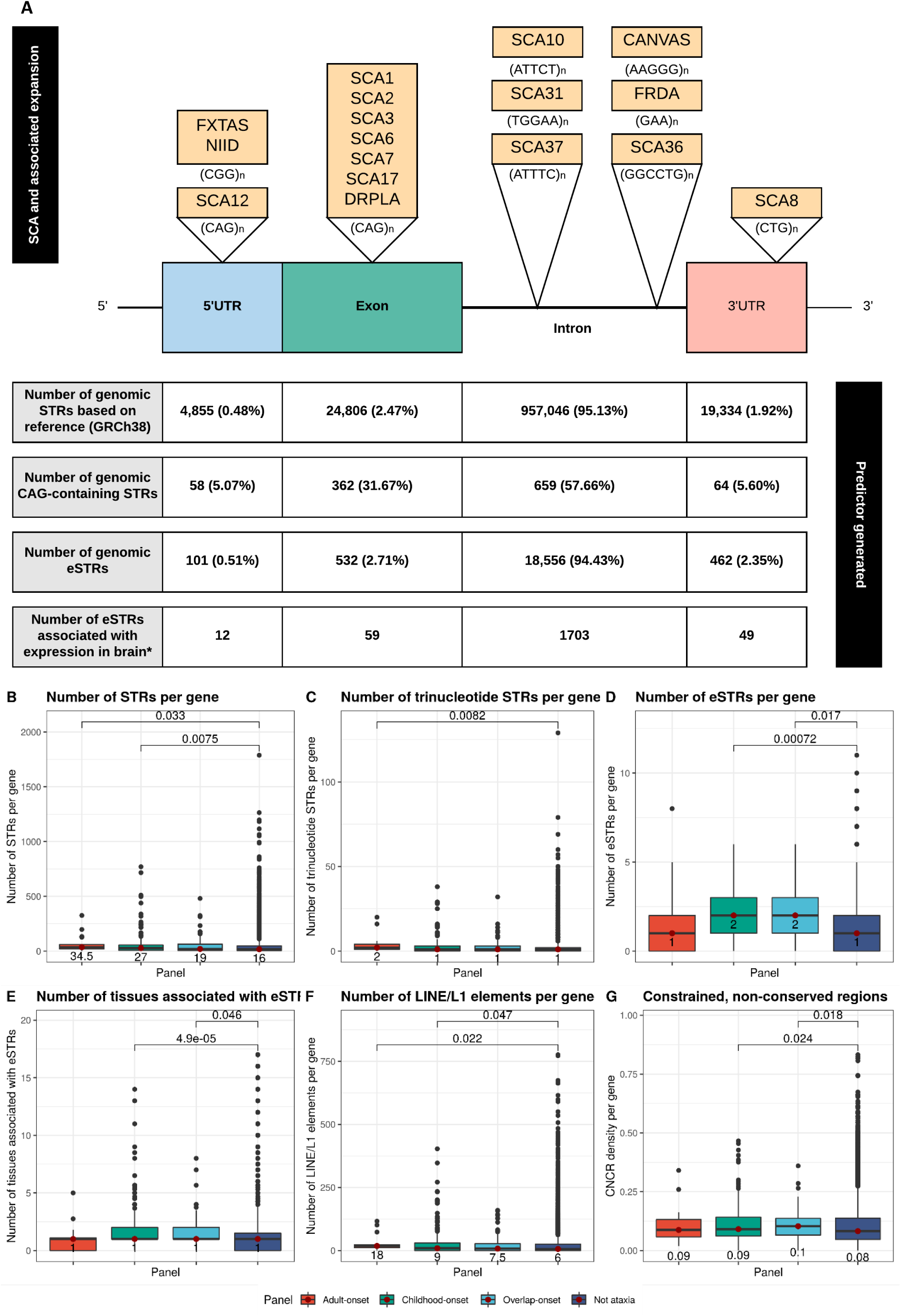
(A) Summary of gene features generated by leveraging information on genomic map of known short tandem repeats (STRs) and expression STRs (eSTRs) with examples of hereditary ataxia associated with pathogenic STRs. The top part of the figure illustrates the location of repeat expansions within spinocerebellar ataxia (SCA) and other ataxias: Fragile X-associated tremor-ataxia syndrome (FXTAS); neuronal intranuclear inclusion disease (NIID); dentatorubral-pallidoluysian atrophy (DRPLA); cerebellar ataxia, neuropathy, and vestibular areflexia syndrome (CANVAS) and Friedreich’s ataxia (FRDA). The bottom part of the figure shows locations of genomic STRs, number of genomic CAG- containing STRs are taken the HipSTR package (Willems *et al.* Nature Methods 2017)^27^. Number of genomic eSTRs are based on analyses from Fotsing *et al*. Nature Genetics 2019.^28^ These eSTRs are taken from those within the top 28,375 eSTRs associated with a high CAVIAR (CAusal Variants Identification in Associated Regions) score for posterior probability of causality when fine-mapped against top 100 nearby SNPs. *Number of eSTRs associated with expression in brain is also derived from this work.^28^ The total number of STRs/ eSTRs is presented with the percentage of overall intragenic location for each STR in parentheses. **(B) Comparison of the number of STRs within each gene (as defined within the HipSTR package) across the four gene lists. (C) Comparison of the number of trinucleotide STRs for each gene across the gene lists. (D) Comparison of the number of eSTRs per gene across the gene lists. (E) Comparison of the number of tissues in which eSTRs affect gene expression across the gene lists. (F) Comparison of the number of LINE/L1 elements per gene (as defined by RepeatMasker) across the gene lists. (G) Comparison of the density of constrained, non-conserved regions (CNCR) per gene across the gene lists.** The CNCR density of a gene reflects the proportion of gene length that is covered by regions fulfilling criteria for constrained but not conserved sequences, such that a density of 1 signifies that the entire gene fulfils criteria for CNCRs. CNCRs are taken from Chen e*t al.* Nature Comms 2021^37^ and reflect the regions of the genome likely to be more human-lineage-specific. Only significant Wilcoxon rank sum p-values (<0.05) are given for pairwise comparisons above the square brackets. The numbers below the red dots in the box and whisker plots represent the median values for that genic feature.

Using RepeatMasker’s library of repetitive elements,^29, 30^ we identified differences in the density of other interspersed repetitive elements between the gene lists. Interestingly, both adult- and childhood-onset genes had a higher number of LINE/L1 elements per gene than controls genes (**Fig. 3F**), as well as SINE/Alu elements among others (**Supplementary Fig. 2**). Thus, our findings demonstrated that ataxia genes are structurally more complex than other genes and this extended to STRs and other repetitive elements. Unexpectedly, these findings were relevant to genes causing childhood- as well as adult-onset ataxia.

### Population-based genetic variation differentiates ataxia genes

Next, we used population-based measures of genetic variation to analyse ataxia genes. As would be expected, the findings largely reflected known differences in inheritance patterns (**Supplementary Fig. 3**). For example, the probability of a gene being intolerant of homozygous and missense variants (gnomadpMiss), but not heterozygous LoF variants from gnomAD data (gnomadpRec),^31^ was significantly higher for childhood-onset genes compared to control genes (**Supplementary Fig. 3**). We also found that CNCR density, a measure of the proportion of human-lineage-specific elements within a gene,^37^ was higher for both childhood- and overlap-onset compared with non-ataxia genes (*P=*0.024, 0.018 respectively) suggesting a higher density of sequences important in human-specific evolution associated with known ataxia genes (**Fig. 3G**).

### Transcriptomic signatures of ataxia genes

Given the phenotypic variability of hereditary ataxia, we explored the tissue- and cell type-specificity of disease genes, as well as their expression over time. The latter was performed using publicly-available temporal expression data generated by RNA-sequencing of human organ development.^38^ We found that there was a significantly higher developmentally-dynamic pleiotropy index representing genes with more repressed temporal expression only within the cerebellum within childhood-onset ataxia genes compared to control genes (*P*=0.042) suggesting timing in expression in earlier development is important (**Fig. 4A**).

**Figure 4.**
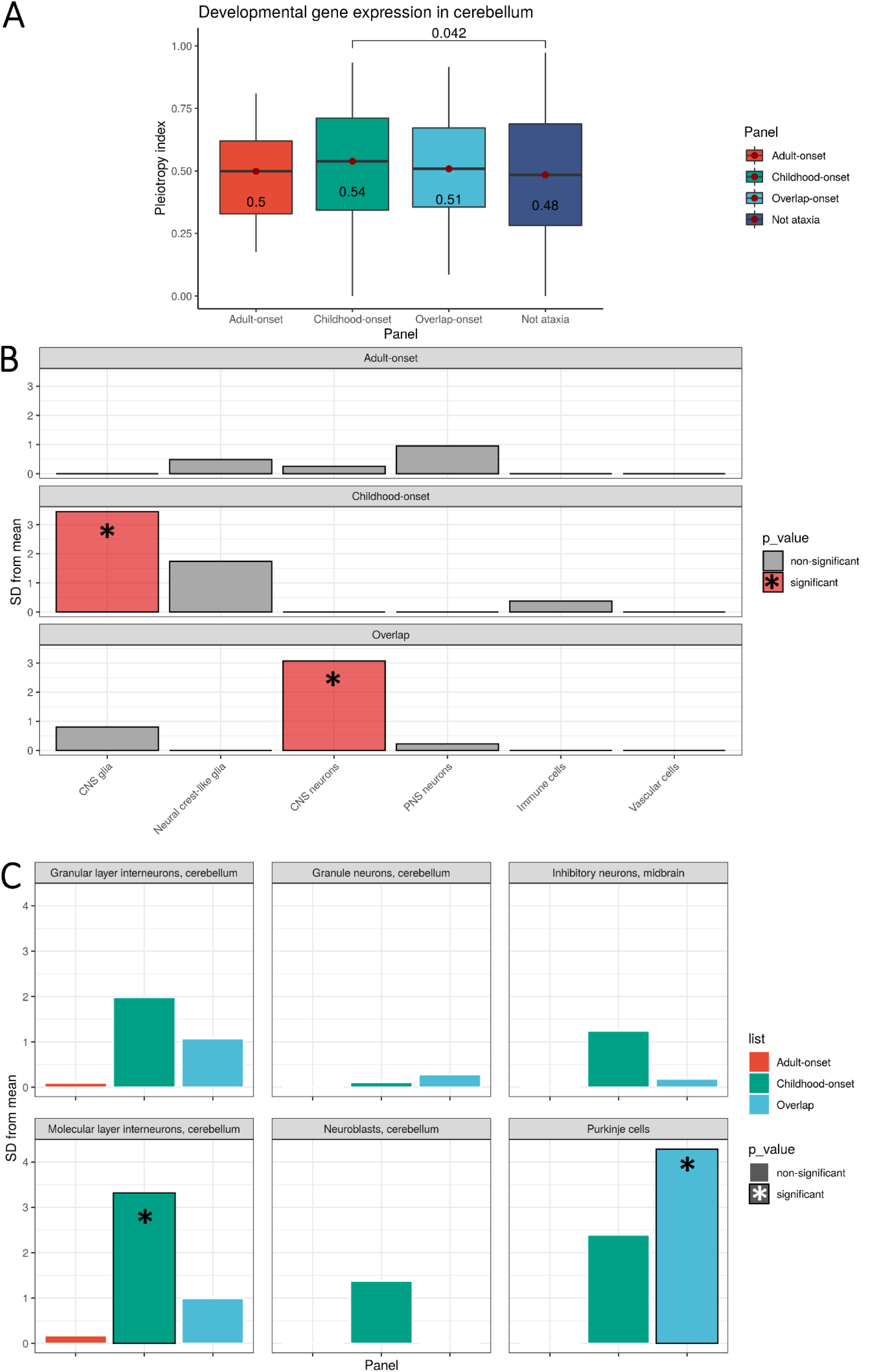
(A) Comparison of markers of dynamic gene expression. Comparison of dynamic specificity indices (where 1 represents repressed temporal expression) in the cerebellum across different gene sets. Only significant Wilcoxon rank sum p-values (<0.05) are given for pairwise comparisons above the square brackets. The numbers below the red dots in the box and whisker plots represent the median values for that genic feature**. Expression-weighted cell-type enrichment (EWCE) results showing significantly- enriched cell-type-specific expression across two levels of cell information. (B) Enrichment of ataxia-associated genes (three sets of different ages of onset) in cell types from mouse single-cell RNA-sequencing data was determined using EWCE.** Standard deviations (SD) from the mean indicate the distance of the mean expression of the target list from the mean expression of the bootstrap replicates. Significance at *P*<0.05 after correction for multiple testing with the Benjamini-Hochberg method over all cell types and the three gene panels was used. CNS refers to central nervous system and PNS refers to the peripheral nervous system. **(C) Enrichment of ataxia-associated genes within cerebellar-specific cell types of the Karolinska dataset are shown with significant p-values noted by an asterix and column outline.**

Next, we used measures of tissue-specific expression and co-expression derived from bulk RNA-sequencing data from 47 human tissues included in GTEx^25^, and found significant differences in tissue-specific expression between the three ataxia gene lists (**Supplementary Fig. 4**). Cerebellar-specific expression appeared to be most associated with the overlap-onset genes but was also an important feature of childhood-onset ataxia, with no statistically significant difference in the proportion of genes with cerebellar tissue-specific expression between the two groups. Similarly, measures of gene co-expression (module membership) highlighted brain-specific tissue co-expression but did not demonstrate significant differences between disease gene sets (**Supplementary Table 5**).

Using EWCE,^41^ together with data on single-cell gene expression profiling of both mouse CNS and non-CNS tissue,^40^ we studied the cell-type-specific expression of ataxia genes. Using all cell types, we demonstrated significant enrichment of CNS glia-specific expression in childhood-onset genes (FDR *P*<1x10^-7^) and CNS neuron-specific expression within overlap-onset genes (FDR *P*=0.018) (**Fig. 4B**). Focusing specifically on cerebellar cell types, we found that childhood-onset genes exhibited cell-type-specific expression within molecular layer interneurons (FDR *P*=0.036) and overlap-onset ataxia genes showed cell-type-specific expression within cerebellar Purkinje cells (FDR *P*=0.018) (**Fig. 4C**). Of interest, childhood-onset genes with the highest cerebellar molecular layer interneuron cell-type expression were associated with ataxia syndromes manifesting partially or fully with seizures (e.g., *KCNA1* and *RORA*). In contrast, overlap-onset genes driving expression in Purkinje cells were associated with ataxia-predominant syndromes including *ITPR1* and *CACNA1G* (**Supplementary Fig. 5**).

### Childhood-onset ataxia genes generate protein products required for viability

Since gene expression levels are often poorly correlated with protein abundance and need not reflect protein-protein interactions,^53^ we assessed all disease genes using the STRING and GO databases.^43, 44^ Consistent with the association of childhood-onset genes with multi- system disorders, we saw a statistically significant enrichment of GO terms associated with glycosylation pathways (e.g., GO:0009101 glycoprotein biosynthetic process) and cilia ontologies (GO:0005929 cilium) (**Fig. 5A**). In contrast, overlap-onset genes were enriched for nervous system-associated terms (e.g., GO:0043005 neuron projection) and ion channel biological processes (GO:0098662 inorganic cation transmembrane transport) (**Fig. 5B**). Adult-onset ataxia genes revealed no significantly enriched GO terms, likely reflecting the small size of this gene list. Given that not all molecular and biological processes are captured by GO, we also analysed the number of protein-protein interactions per gene using data provided by the STRING database. This demonstrated a significantly higher number of protein-protein interactions amongst genes in overlap-onset ataxia than those causing childhood-onset ataxia (*P*=0.018).

**Figure 5.**
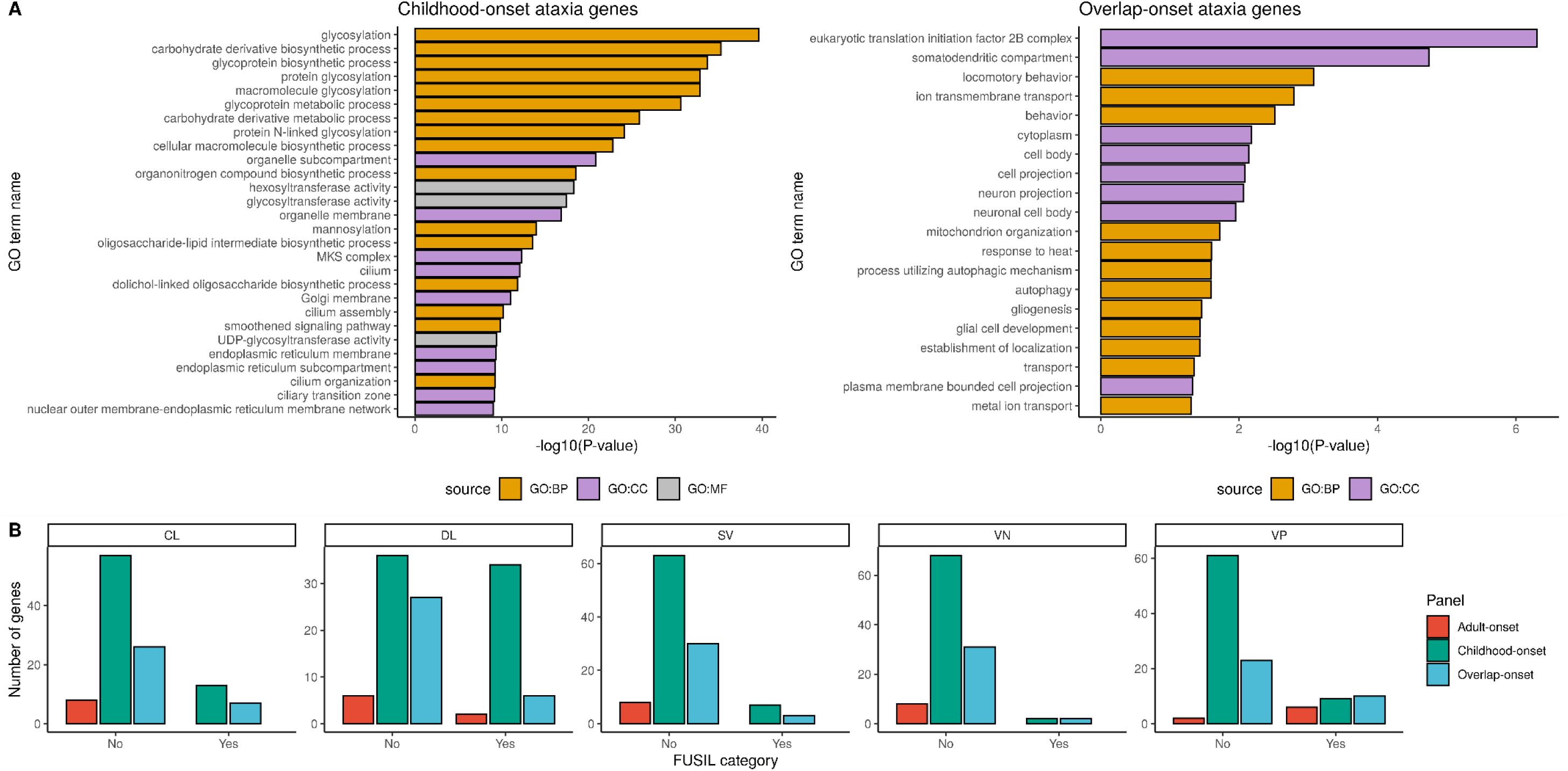
(A) Enriched gene ontology (GO) terms for childhood-onset hereditary ataxia genes (top 25 shown only) and for overlap-onset ataxia genes with associated g:SCS- corrected p-values from gene set analysis. The source depicts the GO of the biological domain with respect to three aspects: BP = biological process, CC = cellular component, MF = molecular function. **(B) Barplots of the number of genes across gene panels are shown for each Full Spectrum of Intolerance to Loss-of-function (FUSIL) category**^46^: CL = cellular lethal. DL = developmental lethal, SV = subviable, VP = viable with phenotypic abnormalities, VN = viable with normal phenotype. “Yes” refers to genes that fulfil criteria for that particular FUSIL category. “No” refers to genes that do no fulfil criteria for that particular FUSIL category.

We extended our analysis to consider functional assessments of gene-protein products by using FUSIL categorisation.^46^ FUSIL classifies genes based on cross-species integrated measures of essentiality.^46^ When comparing childhood- with overlap-onset genes, we saw a significantly higher proportion of genes within the VP category (denoting genes where LoF mutations are viable, but with an abnormal phenotype in mice) in the overlap-onset group (*P*=0.024) (**Fig. 5C**). However, we noted a significantly higher proportion of genes within the CL genes, where LoF mutations cause cellular lethality; *P*=2.467×10^-4^, and DL category genes, where LoF mutations cause developmental lethality; *P*=1.896×10^-17^ amongst childhood-onset genes compared with non-ataxia genes. This reflects the developmental importance of childhood-onset genes.

### A modified testing strategy for variants and STRs in known ataxia genes could improve diagnostic yield

Although we found significant differences in some genic properties across the ataxia gene sets, there were also surprising commonalities, as exemplified by the high density of non-pathogenic STRs in both childhood- and adult-onset genes. To summarise data across all 294 genic features analysed and account for redundancies in correlation between features (**Supplementary Fig. 7**), we used recursive feature elimination to identify the 84 most relevant annotations (**Supplementary Table 6**). Using these salient features, we found that while there were small clusters of childhood- and overlap-onset genes, most ataxia genes did not cluster and adult-onset genes showed no distinct classification on UMAP (**Fig. 6A, Supplementary Fig. 6**). This suggested that adult-onset genes are not easily distinguishable from childhood-onset genes using the genic features assessed, and that the age-of-onset division does not appear to be reflected in functional genomic annotation. This finding has important implications for genetic testing strategies. More specifically, it would suggest that genes currently considered to cause childhood-onset ataxia could be expected to also cause adult-onset disease, and STR expansion may be a more common pathogenic mechanism than expected, potentially operating in combination with known pathogenic variants.

**Figure 6.**
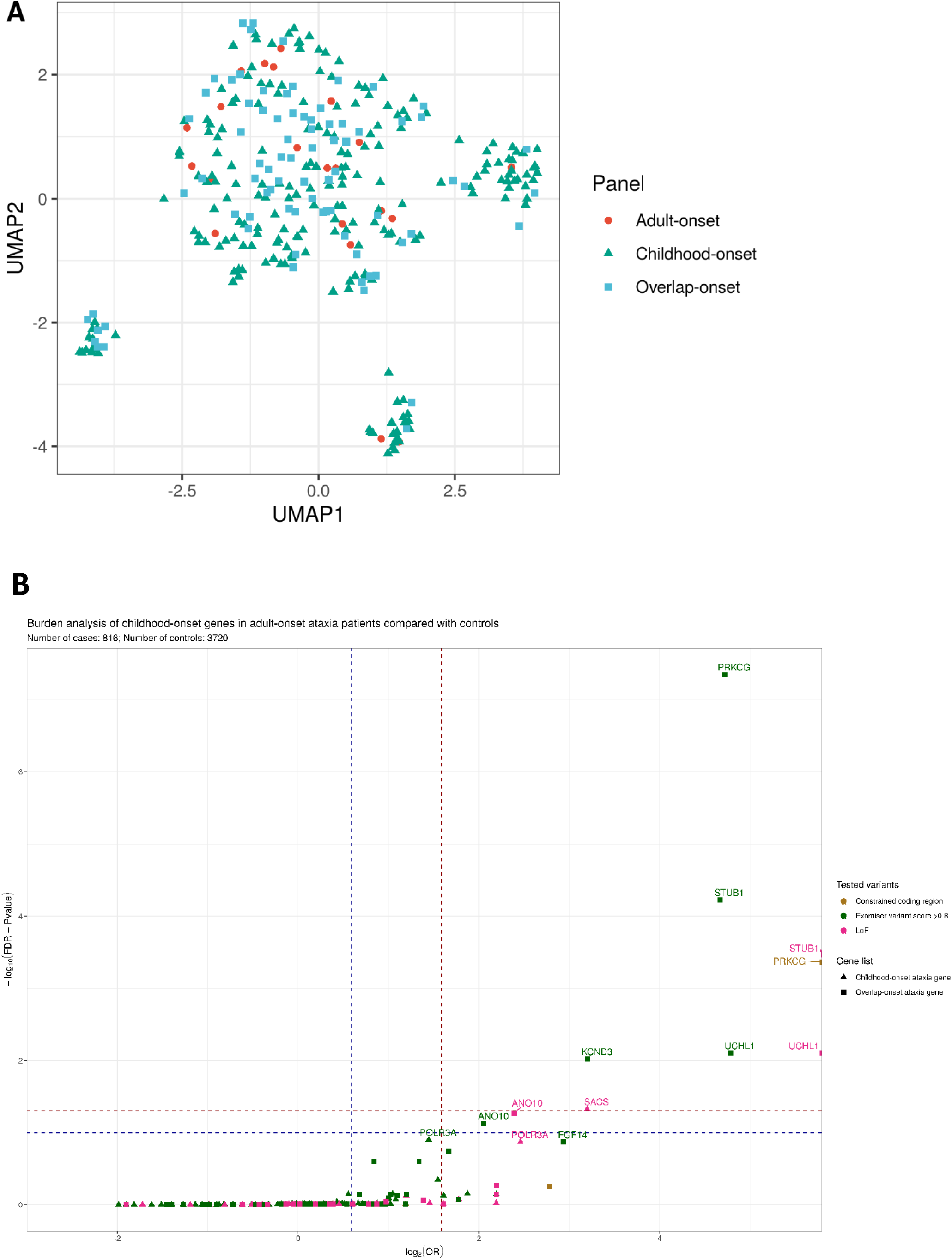
(**A**) **UMAP of all ataxia genes partitioned by age-of-onset using 84 selected genic features from recursive feature elimination.** Results are for each of the three gene panels are shown in Supplementary Figure 6. (**B**) **Volcano plot depicting results from rare variant burden analysis using 100,000 Genomes Project participants.** In this gene-based burden testing analysis, we assessed the number of adult-onset ataxia patients carrying variants in childhood-onset genes filtered for rare variants within constrained coding regions, or with an Exomiser score of >0.8 to indicate likely pathogenicity, or loss-of-function (LoF) variants. We also tested this burden of rare variants in overlap-onset ataxia genes which are expected to be significantly enriched within adult-onset patients. The Odds Ratio (OR) is the odds of enrichment of a variant in cases over controls (defined within Methods). Benjamini- Hochberg method was used to correct for multiple testing; an overall FDR-adjusted *P* of 0.05 (horizontal dashed dark red line) was used for claiming significant gene-disease associations taking into account the total number of case-control gene burden tests under all four scenarios analysed. The horizontal dashed blue line represents an FDR *P* of 0.10. The vertical dashed blue line represents an OR of 1.5 and the dark red dashed vertical line represents an OR of 3.

We explicitly tested the first of these two hypotheses by measuring the burden of potentially pathogenic variants among childhood-onset genes in adult-onset ataxia patients recruited to the 100,000 Genomes Project and vice versa. Interestingly, this demonstrated a significantly higher burden of rare potentially pathogenic LoF variants in the typically childhood-onset ataxia gene *SACS* amongst individuals with adult-onset ataxia (OR 9.178, FDR *P*=0.047) (**Fig. 6B**) (**Supplementary Table 7**). Furthermore, we saw a trend for increased burden of variants in childhood-onset gene *POLR3A* (FDR *P=*0.126 and 0.135 for LoF and Exomiser score >0.8 variants respectively) within adult-onset patients (**Fig. 6B**). This was reflected on interrogation of the clinical records and the final diagnostic-grade genetic reports of ataxia patients within the 100,000 Genomes Project. We identified eight cases where despite the patient presenting with adult-onset disease (range 22 to 47 years of age), diagnoses were made based on a pathogenic variant typically associated with childhood-onset ataxia (in *POLR3A, SACS, PMM2, WFS1*) (**Supplementary Table 8**). This suggests that we may be missing diagnoses in adult patients by not screening for genes typically described to be associated with childhood-onset disorders. Furthermore, an *a priori* assumption of the age-of-onset of an associated gene may cause bias when assessing the variant pathogenicity, especially if the existing described cases are rare.

Furthermore, we assessed the potential importance of STR expansion across a wider set of ataxia genes. More specifically, given the high STR density within ataxia genes (mean of 59.5 STRs per adult-onset gene and 55.8 STRs per childhood-onset gene), we investigated whether ataxia genes not currently thought to cause disease through repeat expansions had higher repeat sizes amongst individuals with ataxia. To assess this, we studied all 191 naturally-occurring 5’UTR and exonic STRs located in 100 genes, of which 116 STRs were classified within genes of typical childhood-onset. We then compared the distribution of STR sizes in ataxia cases (either childhood- or adult-onset) and controls recruited in the 100,000 Genomes Project. Using this approach, we found a trend for a higher maximum number of repeats in the top 1% of repeat sizes in patients presenting with both childhood- and adult-onset ataxia compared with controls (**Fig. 7**). This demonstrated that expansions of apparently benign STRs are associated with disease as evidenced by the trend for higher repeat size in cases over controls even at naturally-occurring STRs. While this does not definitively demonstrate the pathogenicity of specific STRs, it suggests that these STRs could be contributing to disease susceptibility potentially by operating as modifiers of disease risk. Consequently, screening for repeat expansions at STR sites in known ataxia genes should be prioritised in unsolved cohorts.

**Figure 7.**
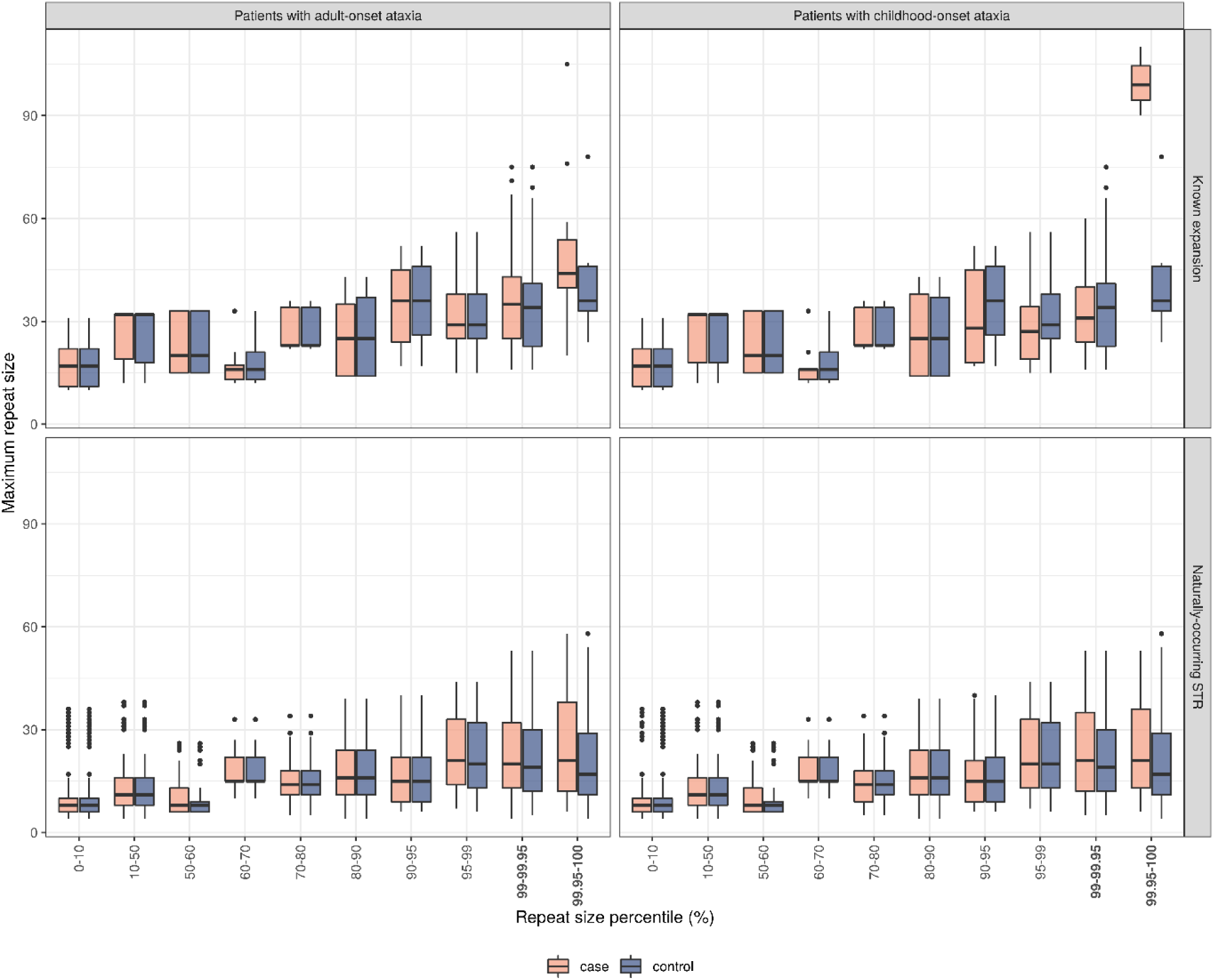
Maximum allelic repeat sizes estimated using ExpansionHunter at STR loci. annotated by HipSTR reference database in adult patients presenting with ataxia (*N* = 1781) and patients presenting with childhood-onset ataxia (*N* = 553) compared with controls (*N* = 6387) defined as unrelated non-neurological probands recruited under the Rare Disease arm of the 100,000 Genomes Project. The repeat sizes were estimated across STRs in which repeat expansions are known to cause ataxia (“Known expansion”) and across naturally- occurring STRs, not currently known to be associated with disease.

## Discussion

In this study, we used 294 functional genomic annotations to study genes causally linked to hereditary ataxia, with the aim of identifying commonalities and differences in gene properties. We provide evidence to show that first, there is an unexpectedly high STR density within childhood-onset ataxia genes suggesting that we may be missing pathogenic repeat expansions as a disease mechanism within this cohort. Secondly, adult-onset ataxia genes cannot be easily distinguished from those causing childhood-onset disease when all genic features are considered, suggesting a common underlying biology. Thirdly and most importantly, diagnostic yield for hereditary ataxia could be improved by using a common screening gene panel and by analysing STRs in existing ataxia genes not known to harbour pathogenic expansions as demonstrated using WGS data from individuals with ataxia in the 100,000 Genomes Project.

By applying a systems biology approach without *a priori* assumptions, we found that genes associated with hereditary ataxia have many common characteristics. Genic features such as increased transcript count did not differentiate between childhood- and adult-onset ataxia, but rather distinguished between ataxia and control genes. Unexpectedly, even STR-based measures of genic complexity were unable to distinguish between adult- and childhood-onset ataxia, as we found a higher density of STRs in genes that currently harbour no known pathogenic repeats, including in childhood-onset ataxia genes. Furthermore, the higher eSTR density within childhood-onset ataxia genes highlighted a potential regulatory role for these elements in modulating gene expression and disease severity. Thus, our analysis provides support for the utility of screening for pathogenic repeat expansions causing hereditary ataxia at all ages of onset. With this in mind, we noted that five previously-undiagnosed children presenting with ataxia in the 100,000 Genomes Project were found to have repeat expansion disorders typically associated with adult-onset disease, for which they had not been initially screened.^16^ Similarly, we provided data to suggest that even STRs not known to be associated with pathogenic repeat expansions tended to have higher repeat sizes in individuals with both adult- and childhood-onset ataxia as compared to controls. This highlights naturally-occurring STRs within known ataxia genes as candidates for screening in an unsolved cohort. Intriguingly, we also found a higher density of other repetitive elements such as LINE/L1 in both adult- and childhood-onset ataxia genes than controls. This finding is of interest in highlighting the potential role of LINE/L1 dysregulation in pathogenesis, as found by a recent RNA-sequencing study that demonstrated cerebellar LINE/L1 activation in driving ataxia phenotype in mouse models.^54^

Our analyses highlighted not only the potential for common pathogenic mechanisms for disease across hereditary ataxia, but also common biology. Intriguingly, we found no clear separation of genes causing childhood- and adult-onset ataxia using UMAP to visualise genes based on a recursively-selected set of features. Most strikingly, adult-onset ataxia genes were highly scattered suggesting that there is a spectrum of disease and that genes causing childhood-onset ataxia have the potential to cause adult-onset disease. We found support for this hypothesis through the demonstration of a significantly higher burden of rare potentially pathogenic variants in the conventionally childhood-onset genes defined within OMIM^21^: *SACS* and *POLR3A* amongst individuals with adult-onset ataxia enrolled in the 100,000 Genomes Project. Furthermore, we recognise that for *SACS*, clinical reports exist of adult disease presentations.^55^ On review of final diagnostic genetic reports, we found eight patients who presented with adult-onset ataxia being diagnosed with variants typically associated with childhood-onset disease within the 100,000 Genomes Project. However, there has not been a systematic approach to gene screening, and these findings would suggest that we could be missing diagnoses in adult-onset patients by either not screening for, or not prioritising pathogenic variants that have been typically described in children.

While our study has largely uncovered significant commonalties across the childhood- and adult-onset ataxia, there were also differences. Given the cellular complexity of the cerebellum, a region that harbours more than half of all brain neurons,^56^ this is unsurprising, and is already supported by evidence that different cerebellar cell types are affected in different forms of ataxia.^57^ Consistent with previous gene expression analyses^58^ and the cerebellum being the most frequent site of neurodegeneration on neuropathological examination of ataxia,^57^ we found that ataxia genes exhibited cerebellar-specific expression and co-expression. Interestingly, we demonstrated a preferential CNS glial enrichment within childhood-onset genes compared to a CNS neuronal enrichment within overlap-onset genes. We found that CNS glial enrichment was driven by ciliopathy genes, which are known to affect early brain development, mediated in part through radial glial progenitor cells.^59^ This finding was also reflected within GO enrichment analyses, showing that childhood-onset genes are associated with glycosylation and cilia pathways. As would be expected, childhood-onset genes not only exhibited dynamically-repressed expression in the cerebellum when compared with control genes,^38^ but a higher proportion were also classified as cellularly or developmentally lethal. Genes causing both adult- and childhood-onset ataxia exhibited Purkinje cell-type-specific expression, driven by genes associated with ‘pure’ ataxia syndromes; a finding supported by previous analyses using mouse^60^ and human transcriptomics.^58^ The expression of childhood-onset ataxia genes was also enriched within inhibitory GABAergic molecular layer interneurons,^61^ driven by genes associated with ataxia-epilepsy syndromes. This supports additional function for these interneurons in epileptogenesis, mirroring the role of dentate basket cells in temporal lobe epilepsy.^62^

Although this study highlighted key biological information, such as the contribution of particular cell types, and the potential importance of specific pathogenic processes such as STR expansions, our analyses were limited by the quality and availability of existing functional genomic annotation. For example, information regarding dynamic gene expression taken from human tissues^38^ is limited by the resolution of bulk RNA-sequencing data as not all genes are successfully sequenced at all timepoints, thus dynamic gene expression can only be quantified in the limited number of genes. Likewise, we are also limited by the accuracy of input gene lists and age-of-onset classification which changes regularly with new cases of hereditary ataxia described. We attempted to overcome this problem by using a range of different resources across the four main gene feature categories and disease gene panels.

In summary, this study suggests that childhood- and adult-onset ataxia exist across a spectrum of disease rather than as distinct entities; a finding which would be hard to generate from clinical experience given that there are many hereditary ataxia genes each accounting for a very small number of cases. This core observation has important clinical implications for the classification of hereditary ataxia by age-of-onset. But, most importantly, it suggests that the diagnostic rate for hereditary ataxia would be expected to increase simply by removing the age-of-onset partition, and through modified screening for repeat expansions in naturally-occurring STRs within known ataxia genes, in effect treating these regions as candidate pathogenic loci.

## Data Availability

All data produced in the present study are available upon reasonable request to the authors.

## Acknowledgements

This research was made possible through access to the data and findings generated by the 100,000 Genomes Project. The 100,000 Genomes Project is managed by Genomics England Limited (a wholly owned company of the Department of Health and Social Care). The 100,000 Genomes Project is funded by the National Institute for Health Research and NHS England. The Wellcome Trust, Cancer Research UK and the Medical Research Council have also funded research infrastructure. The 100,000 Genomes Project uses data provided by patients and other participants collected by the National Health Service as part of their care and support. We thank all participants and healthcare teams at the thirteen NHS Genomic Medicine Centres in England, where around 5,000 multidisciplinary staff enrolled patients to the 100,000 Genomes Project. Patients were also enrolled to the 100,000 Genomes Project from Scotland by the Scottish Genomes Project, and across Wales and Northern Ireland.

ZC was supported by a clinical research fellowship from the Leonard Wolfson Foundation. AT is a Medical Research Council (MRC) clinician scientist (MR/S006753/1). JH was supported by the UK Dementia Research Institute which receives its funding from DRI Limited, funded by the UK MRC, Alzheimer’s Society and Alzheimer’s Research UK. JH has also been funded by the MRC (MR/N026004/1), Wellcome Trust (202903/Z/16/Z), Dolby Family Fund and National Institute for Health Research University College London Hospitals Biomedical Research Centre. JB is supported through the Science and Technology Agency, Séneca Foundation, CARM, Spain (research project 00007/COVI/20). MR was supported by the UK MRC through the award of a Tenure Track Clinician Scientist Fellowship (MR/N008324/1). The views expressed are those of the authors and not necessarily those of the NHS, the funding bodies or the Department of Health and Social Care.

## Author contributions

ZC and MR conceived and designed the study. ZC curated the gene lists, genic features, analysed the data, drafted the figures and the initial manuscript. ZC, AT, EKG, NW, HH and MR reviewed the clinical data and age-of-onset ataxia gene lists, and provided clinical insight into the data. AT and KI supervised and performed the ExpansionHunter genotyping of STRs within the 100,000 Genomes Project. VC, LV and DS performed gene burden analysis within the 100,000 Genomes Project. ZC, RHR, DZ, ACG, SS and JB curated the genic features, performed EWCE and UMAP. All authors contributed to the critical analysis and revision of the manuscript. HH, JB and MR supervised the project. GERC provided data and findings generated by the 100,000 Genomes Project.

### Supplementary material

Supplementary material is available at *Brain* online.

## Appendix I

### Genomics England Research Consortium

Ambrose J. C. ^1^, Arumugam P.^1^, Baple E. L. ^1^, Bleda M. ^1^, Boardman-Pretty F. ^1,2^, Boissiere J. M. ^1^, Boustred C. R. ^1^, Brittain H.^1^, Caulfield M. J.^1,2^, Chan G. C. ^1^, Craig C. E. H. ^1^, Daugherty L. C. ^1^, de Burca A. ^1^, Devereau, A. ^1^, Elgar G. ^1,2^, Foulger R. E. ^1^, Fowler T. ^1^, Furió-Tarí P. ^1^, Hackett J. M. ^1^, Halai D. ^1^, Hamblin A.^1^, Henderson S.^1,2^, Holman J. E. ^1^, Hubbard T. J. P. ^1^, Ibáñez K.^1,2^, Jackson R. ^1^, Jones L. J. ^1,2^, Kasperaviciute D. ^1,2^, Kayikci M. ^1^, Lahnstein L. ^1^, Lawson K. ^1^, Leigh S. E. A. ^1^, Leong I. U. S. ^1^, Lopez F. J. ^1^, Maleady-Crowe F. ^1^, Mason J. ^1^, McDonagh E. M. ^1,2^, Moutsianas L. ^1,2^, Mueller M. ^1,2^, Murugaesu N. ^1^, Need A. C. ^1,2^, Odhams C. A.^1^, Patch C. ^1,2^, Perez-Gil D. ^1^, Polychronopoulos D. ^1^, Pullinger J. ^1^, Rahim T. ^1^, Rendon A. ^1^, Riesgo-Ferreiro P.^1^, Rogers T. ^1^, Ryten M. ^1^, Savage K. ^1^, Sawant K. ^1^, Scott R. H. ^1^, Siddiq A. ^1^, Sieghart A. ^1^, Smedley D. ^1,2^, Smith K. R. ^1,2^, Sosinsky A. ^1,2^, Spooner W. ^1^, Stevens H. E. ^1^, Stuckey A. ^1^, Sultana R. ^1^, Thomas E. R. A. ^1,2^, Thompson S. R. ^1^, Tregidgo C. ^1^, Tucci A. ^1,2^, Walsh E. ^1^, Watters, S. A. ^1^, Welland M. J. ^1^, Williams E. ^1^, Witkowska K. ^1,2^, Wood S. M. ^1,2^, Zarowiecki M.^1^ .

1. Genomics England, London, UK

2. William Harvey Research Institute, Queen Mary University of London, London, EC1M 6BQ, UK.

## Supplementary Figure Legends

**Supplementary Figure 1.**
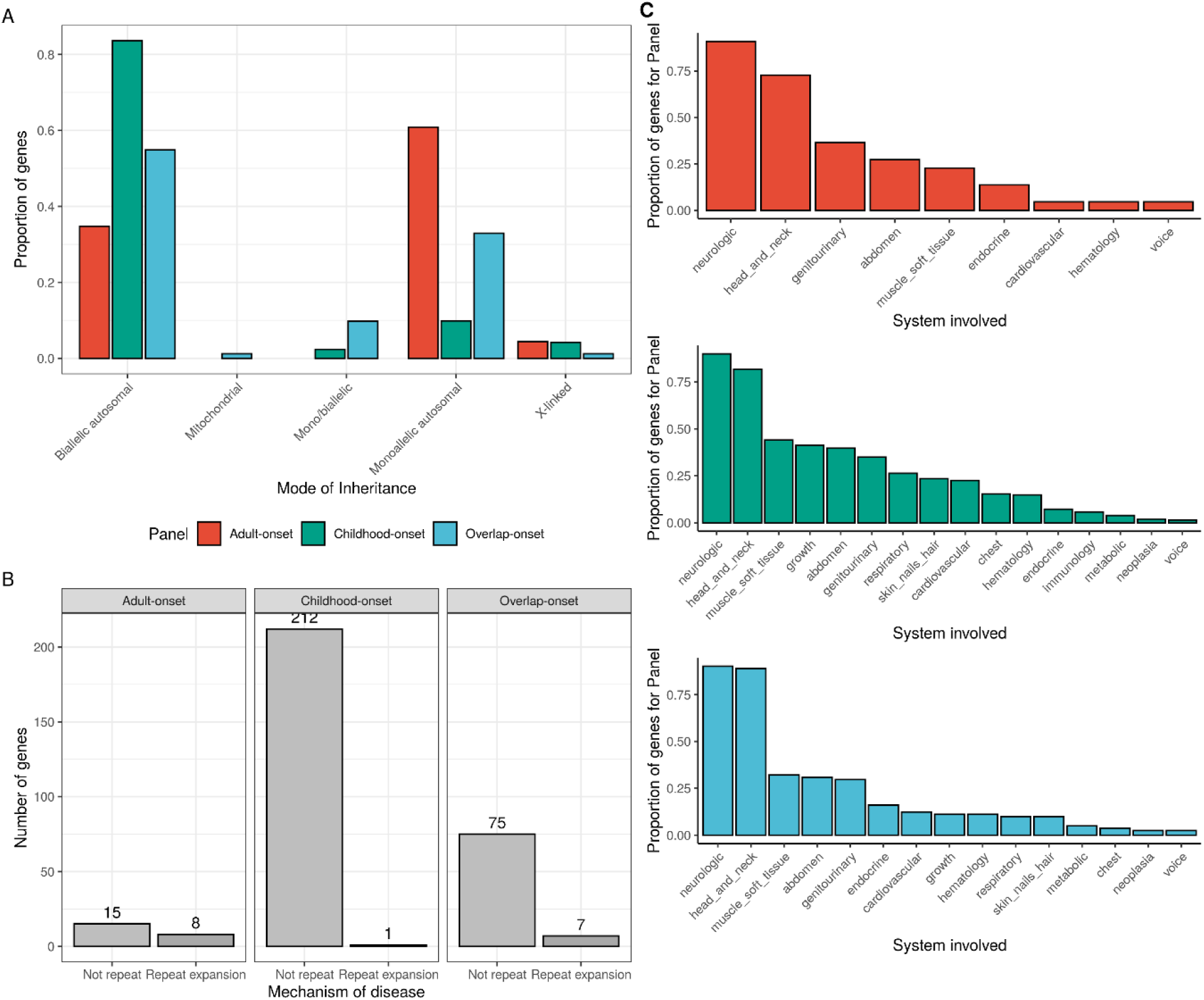
Summary of the differences between genes known to cause ataxias using existing knowledge. (**A**) Proportion of genes with mode of inheritance across adult-onset, childhood-onset and overlap-onset hereditary ataxia genes. (**B**) Number of genes with known repeat expansions causing hereditary ataxia and number of genes not known to be associated with repeat expansion disorder. (**C**) Proportion of genes for that age-of-onset gene panel associated with disease in a particular system of the body as defined by OMIM across adult-, childhood- and overlap-onset ataxia genes.

**Supplementary Figure 2.**
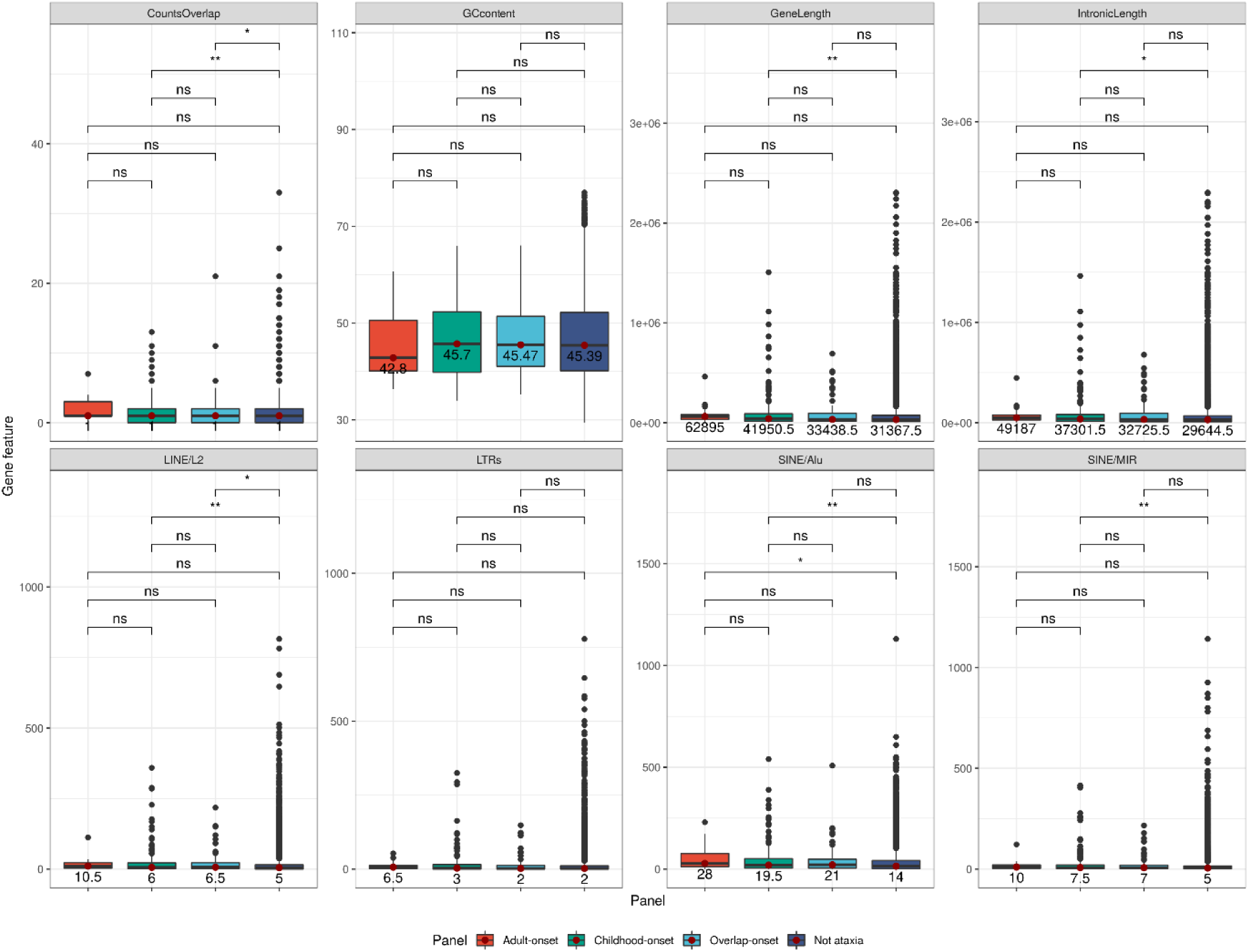
Comparison of genic features capturing information about gene structure and gene complexity. CountsOverlap refers to the number of genes overlapping a gene of interest, GCcontent refers to the percentage GC content of a given gene, GeneLength is the length of each gene defined by its transcription start and end sites given in base pairs, IntronicLength is the total length of annotated introns within a gene. The bottom four panels shown the number of repeat elements per gene of LINE/L2 (long interspersed nuclear elements); LTRs (long terminal repeats), SINE/Alu (short interspersed nuclear elements/Alu sequences); SINE/MIR (SINE/mammalian-wide interspersed repeats) across the four different gene panels. Wilcoxon rank sum p-value comparing the gene lists are shown as follows: ns: p > 0.05; *: p < 0.05; **: p < 0.01; ***: p < 0.001; ****: p < 0.0001. The numbers below the red dots in the box and whisker plots represent the median values for that genic feature.

**Supplementary Figure 3.**
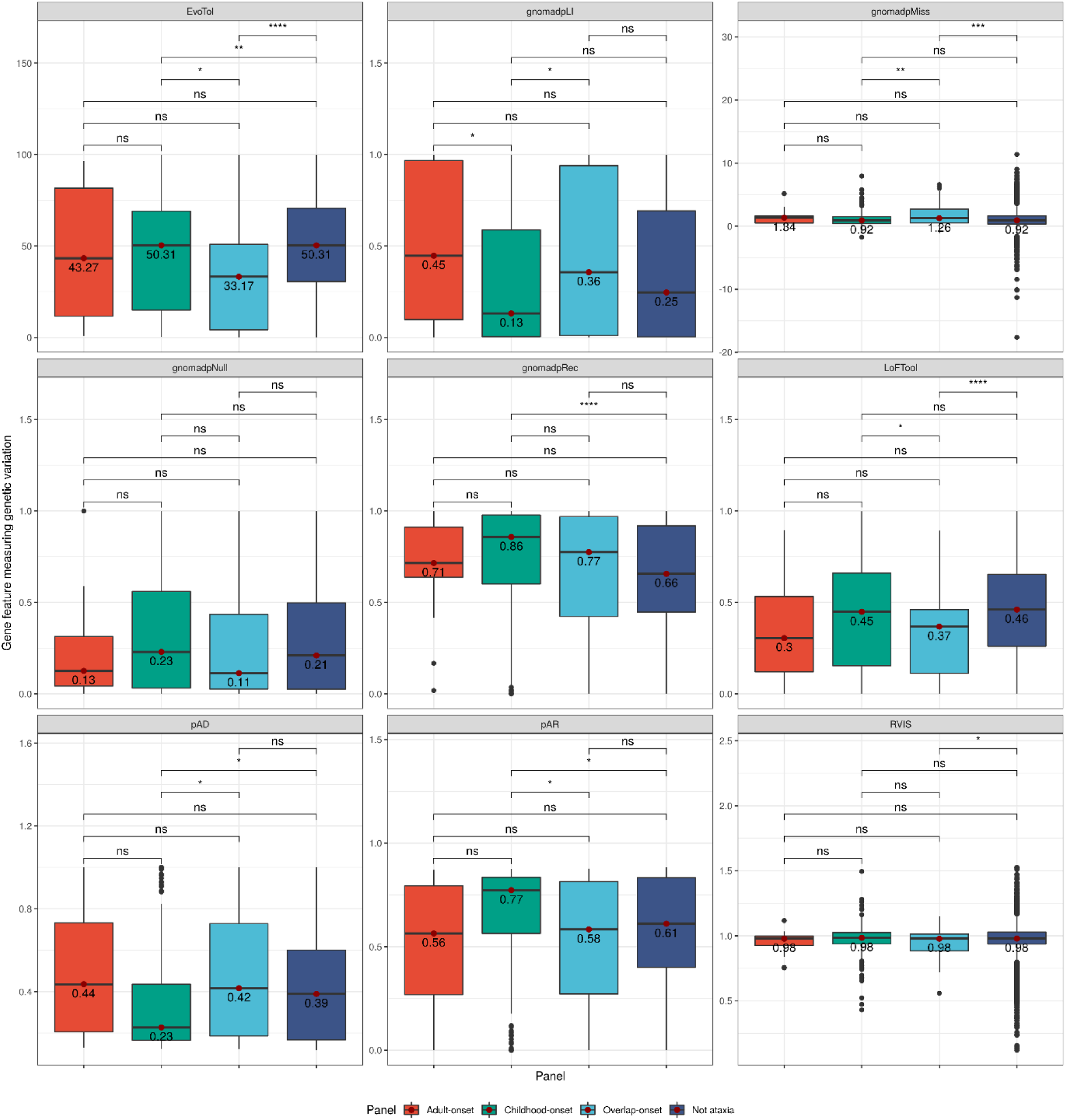
Comparing genic measures of genic variation using data from publicly-available databases. LoFTool (gene intolerance score based on loss-of-function variants); EvoTol (quantifies a gene’s intolerance to mutation using evolutionary conservation of protein sequences); RVIS (intolerance scoring system that assesses whether genes have more or less functional genetic variation than expected based on the apparently neutral variation found in the gene); pAD (probability of autosomal dominant variants); pAR (probability of autosomal recessive variants); gnomadpLI (loss-of-function score from gnomAD: pLI closer to 1 indicates that the gene or transcript cannot tolerate protein-truncating variation); gnomadpRec (probability of being intolerant of homozygous, but not heterozygous loss-of-function variants); gnomadpNull (probability of being intolerant to both homozygous and heterozygous variants); gnomadpMiss (probability of being intolerant of missense variants. Wilcoxon rank-sum p-value comparing the gene lists are shown as follows: ns: p > 0.05; *: p < 0.05; **: p < 0.01; ***: p < 0.001; ****: p < 0.0001. The numbers below the red dots in the box and whisker plots represent the median values for that genic feature.

**Supplementary Figure 4.**
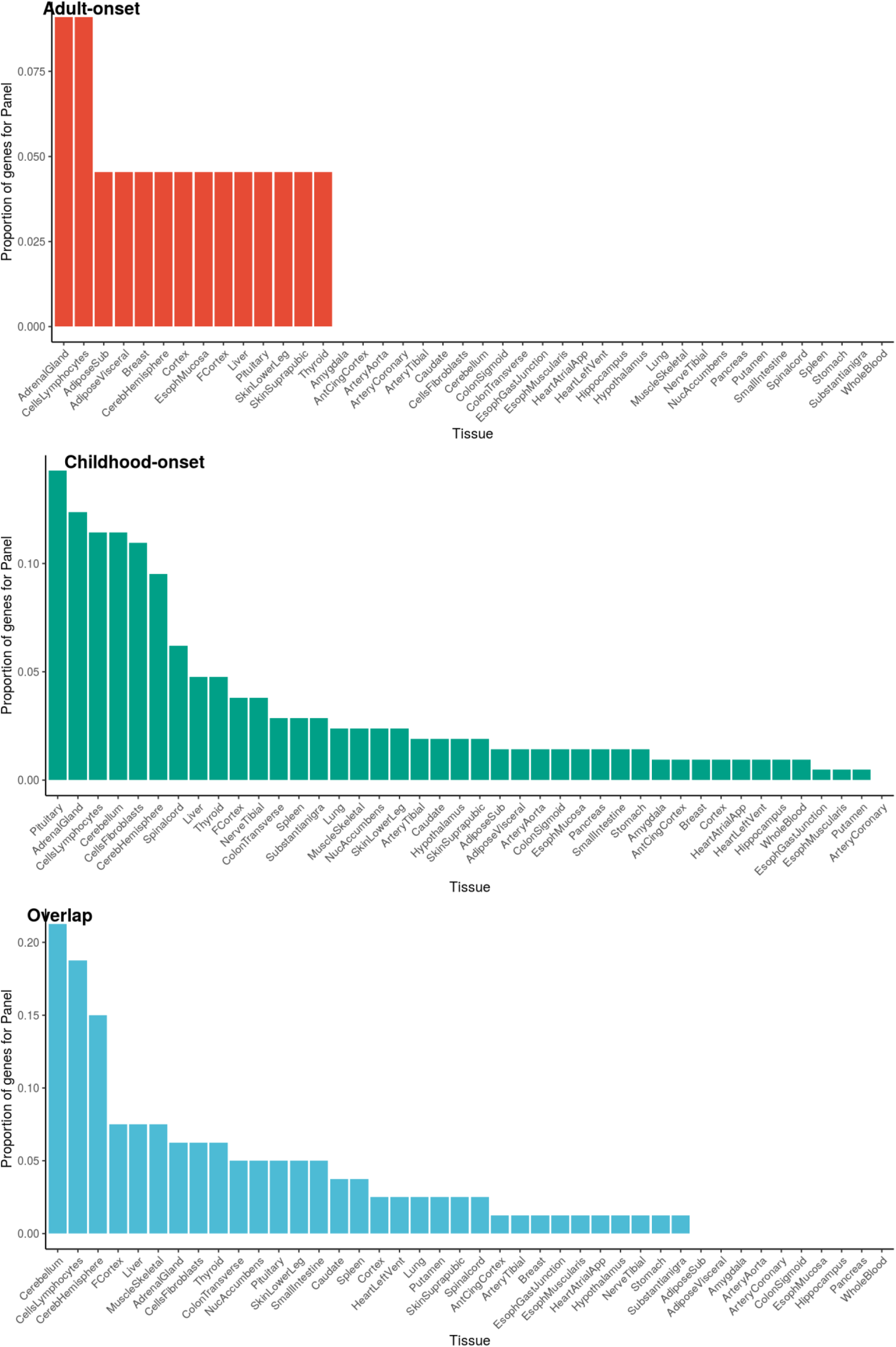
Proportion of genes within each ataxia gene panel partitioned by age-of-onset with tissue-specific expression for each of the 47 Genotype-Tissue Expression Project (GTEx) tissues. Gene expression data were filtered for genes with >0.1 Reads Per Kilobase Million (RPKM) and corrected for batch effects, age, sex and RNA integrity number using ComBat. Residuals of these linear regression models were used to calculate tissue-specific expression. A gene was defined as having tissue-specific expression if expression in that tissue was five-fold higher than the mean across all tissues.

**Supplementary Figure 5.**
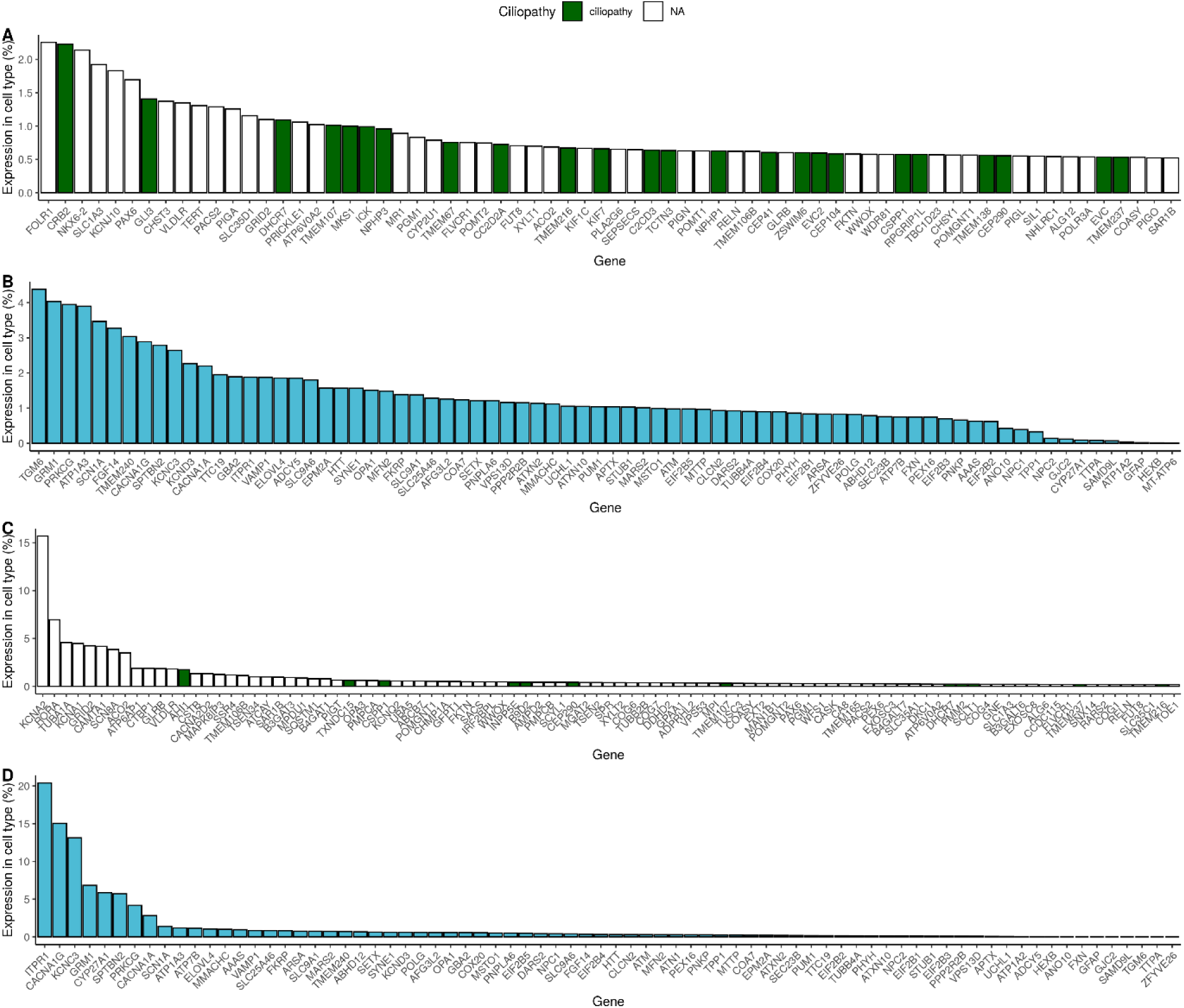
Plot of specificity values for ataxia-associated genes. Specificity values were derived from Skene et al. (2018) who calculated specificity by dividing the mean expression of a gene in one cell type by the mean expression in all cell types. Specificity is the proportion of a gene’s total expression attributable to one cell type, with a value of 0% meaning a gene is not expressed in that cell type and a value of 100% meaning that a gene is only expressed in that cell type. **(A)** Childhood-onset genes within CNS glia (level 2 cell types from the Karolinska single-cell RNA-sequencing) with ciliopathy genes shown in green; **(B)** Overlap-onset genes within CNS neurons (level 2 cell types); **(C)** Childhood-onset genes within cerebellar molecular interneurons (level 3 cell types from the Karolinska superset); (**D)** overlap-onset genes within cerebellar Purkinje cells (level 3).

**Supplementary Figure 6.**
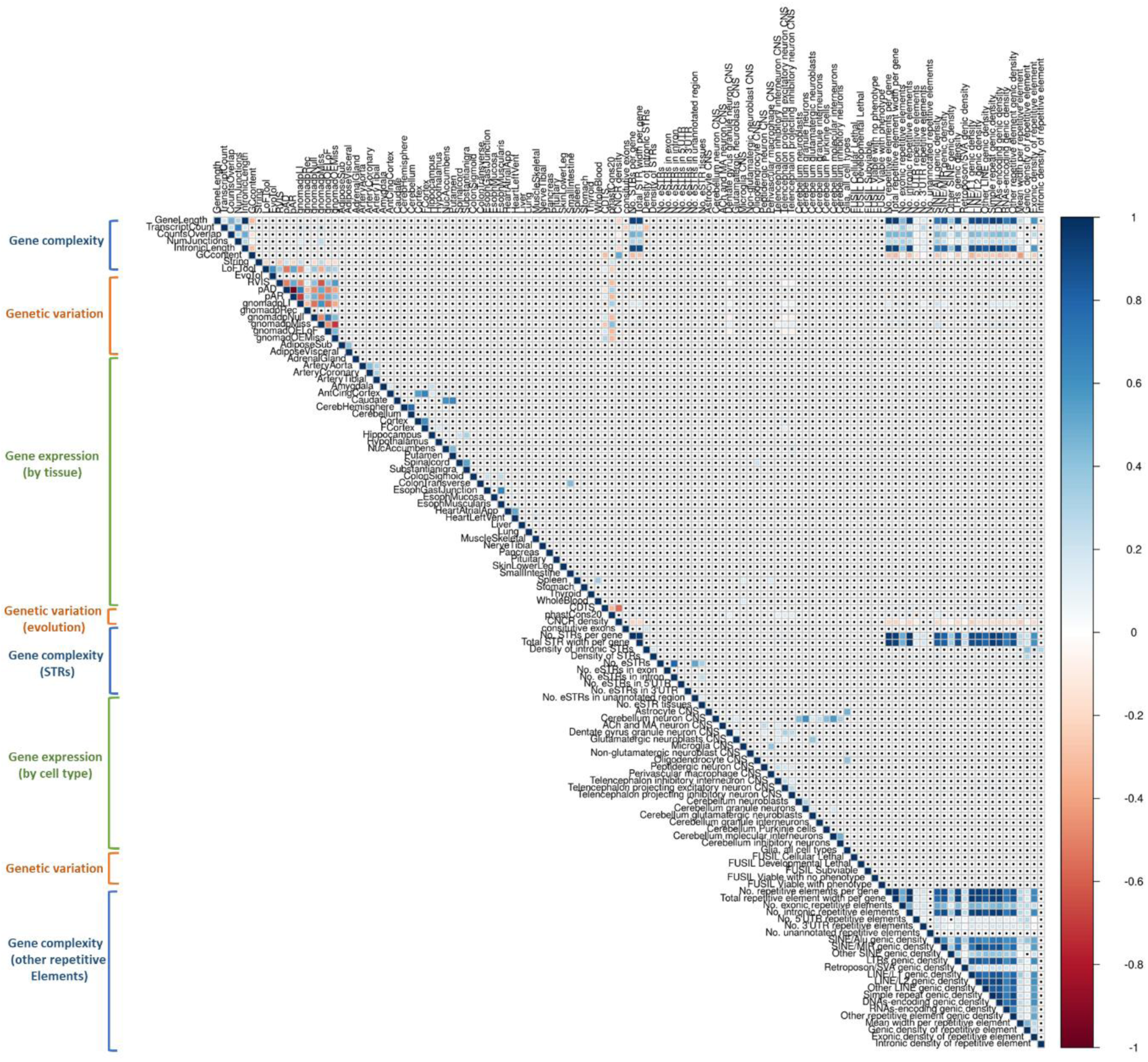
Correlation matrix of the gene features classed by major groups of gene complexity, genetic variation and gene expression, by tissue and cell type. Pearson’s correlation values are represented by the gradient shown. Module membership, adjacency, level 1 cell-type-specific expression are not shown in the plot for ease of data visualisation. Explanation for each individual feature is provided in Supplementary Table 2.

**Supplementary Figure 7.**
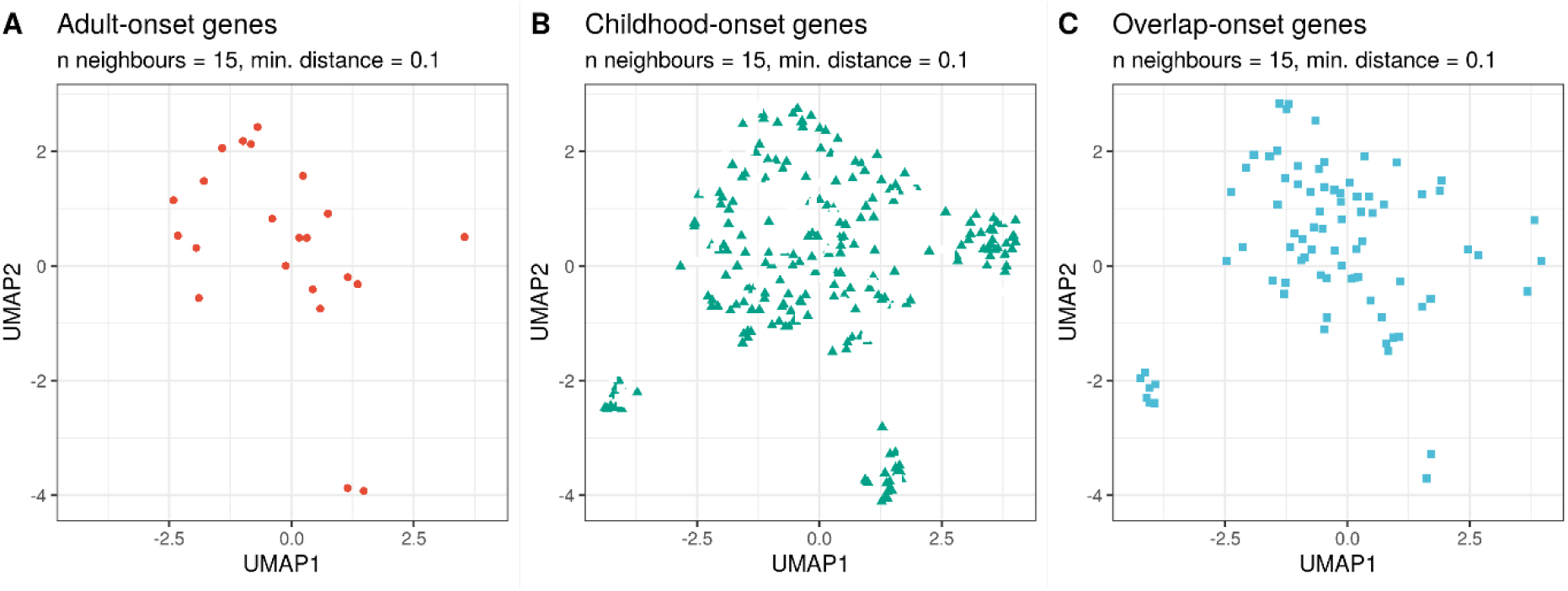
Individual UMAP of ataxia genes partitioned by age-of-onset using 84 selected genic features from recursive feature elimination. **(A)** Adult-onset genes; (**B**) childhood-onset genes; (**C**) Overlap-onset genes.

